# Exploring the Link Between Body Physiology and Cognition: The Role of the Brain and Aging

**DOI:** 10.64898/2026.01.13.26343950

**Authors:** Irina Buianova, Narun Pat

**Affiliations:** Department of Psychology, University of Otago, Dunedin, New Zealand

**Keywords:** cognition, aging, body physiology, neuroimaging, body composition

## Abstract

Epidemiological links between cognition and body physiology in aging are well established, but their strength and drivers remain unclear. Which physiological systems – from body composition to cardiovascular, pulmonary, renal, hepatic, immune, metabolic, and musculoskeletal – best predict cognition, and to what extent are cognition–body associations linked to brain variation across aging? We examined 19 physiological phenotypes alongside three neuroimaging modalities in over 30,000 UK Biobank participants. Machine learning models integrating body measures predicted cognition at *r*=0.4, demonstrating a cognition–body covariation at 16%. Body composition and bone health emerged as the strongest predictors. Notably, 85.1% of cognition–body covariance overlapped with neuroimaging, especially white matter features. Moreover, 71.7% of cognition–age covariance was jointly shared with neuroimaging and physiology, and 96.8% was shared with either brain or body markers, or their overlap. Together, these findings clarify how body physiology and brain structure and function covary with cognitive aging.

## INTRODUCTION

“Mens sana in corpore sano” is a Latin phrase meaning “a healthy mind in a healthy body.” This well-known expression captures the long-recognised link between cognitive and physical health. Empirical research has demonstrated this connection across multiple physiological systems, including body composition, cardiovascular (Pacholko & Iadecola, 2024; Song et al., 2020; Williams et al., 2025), pulmonary (Duggan et al., 2020; Grande et al., 2024; Ma et al., 2025), renal (Bronas et al., 2017; Capasso et al., 2025), hepatic (Filipović et al., 2018; Gao et al., 2024; Zhang et al., 2024), immune (Liu et al., 2024; Shaw et al., 2024; Zou et al., 2025), metabolic (Dimache et al., 2021; Koutsonida et al., 2022; Röhr et al., 2020), and musculoskeletal systems (Huang et al., 2025; Laudisio et al., 2016; Qu et al., 2020).

However, despite extensive evidence linking bodily physiology to cognition in aging, prior work has largely examined isolated physiological systems or pairwise associations rather than integrative, multimodal approaches that jointly characterize body physiology, brain variation, and cognition. Most studies have also focused on associations observed within the same group of participants used to develop the models, rather than evaluating whether system-wide physiological measures considered holistically can predict cognitive functioning in new individuals who were not part of the modelling process (Karlamangla et al., 2014). Consequently, less is known about the relative predictive value of system-wide physiological indicators, how well these relationships generalize to unseen individuals, and the extent to which body–cognition associations overlap with multimodal neuroimaging characteristics. Evaluating the predictive capacity of system-wide physiology will allow us to quantify the overall strength of the cognition–body relationship and identify the physiological systems most strongly linked to cognitive variation.

Equally important is our limited understanding of how and to what extent the cognition–body relationship across multiple physiological systems is reflected in different aspects of the brain, as measured through various neuroimaging modalities (Min et al., 2024). Diffusion-weighted MRI (dwMRI) enables the indirect evaluation of white matter microstructural integrity and organisation (Pierpaoli et al., 1996). Resting-state functional MRI (rsMRI) captures temporal correlations in the blood oxygenation level dependent (BOLD) signal across brain regions (Lee et al., 2013). Structural MRI (sMRI) reflects the anatomy and morphology of grey and white matter (Symms et al., 2004). Several studies have used these modalities to develop predictive markers of cognitive functioning, achieving reasonable performance (Dhamala et al., 2021; Krämer et al., 2024; Rasero et al., 2021; Sripada et al., 2020). Yet, it remains unknown how predictive brain markers of cognitive functioning – whether unimodal, based on individual neuroimaging modalities, or composite, integrating information across multiple modalities – compare with markers derived from body phenotypes, and, more importantly, how much of the cognition–body relationship overlaps with these brain markers. Clarifying this will provide a more comprehensive understanding of the cognition–body relationship by revealing the extent to which brain markers mirror the associations between body physiology and cognition.

The relationship between cognition and body is not static, as cognitive functioning declines with age (Salthouse, 2019). However, it remains poorly understood to what extent physiology across multiple body systems is associated with age-related cognitive variation, and to how much of this link is reflected in the brain. Examining the extent to which body and brain jointly capture the relationship between cognition and age will help determine the potential of system-wide physiology as a predictive marker of cognitive aging and clarify the respective contributions of body and brain across systems and modalities.

Using the UK Biobank, one of the largest datasets combining measures of cognitive performance, body physiology, and neuroimaging, we applied machine learning to predict cognitive functioning from 19 body and 81 brain phenotypes. Cognitive functioning was operationalized as the general factor of cognition (*g*-factor), a latent construct capturing shared variance across multiple cognitive domains and tasks (Jensen, 2000; Salthouse, 2013). Nineteen body phenotypes comprised 317 measures representing overall body composition and seven physiological systems: cardiovascular, pulmonary, renal, hepatic, immune, metabolic, and musculoskeletal. Brain phenotypes included neuroimaging-derived measures from three MRI modalities: dwMRI, rsMRI, and sMRI.

The current study had three aims. First, we used machine learning to assess how well individual body phenotypes and a composite measure integrating all phenotypes predicted cognition in held-out participants not used in the model-building process. We benchmarked these body-based models against unimodal and composite brain-based models, comparing predictive accuracy, feature importance, and performance across body systems. Second, we quantified the extent to which cognition–body covariance overlapped with brain markers, investigating how similarly brain and body measures relate to cognition. Finally, we assessed how much of the cognition–age covariance was shared with composite body and brain measures, examining how similarly brain and body measures and age relate to cognition.

## RESULTS

### Cognition

We derived the *g*-factor from twelve cognitive performance scores using exploratory structural equation modelling within confirmatory factor analysis (ESEM-within-CFA, https://mateuspsi.github.io/esemComp/articles/esem-within-cfa.html). The four-factor structure was confirmed through parallel factor analysis, and construct validity was further established through CFA, supporting the interpretation of both domain-specific and general cognitive abilities (Fig. 1). Supplementary Table S1 lists the cognitive tests included in the study. Supplementary Table S2 presents the loadings of the twelve cognitive performance metrics on latent variables in the ESEM-within-CFA model, and Supplementary Table S3 summarises goodness-of-fit indices for the hierarchical *g*-factor model.

**Figure 1.**
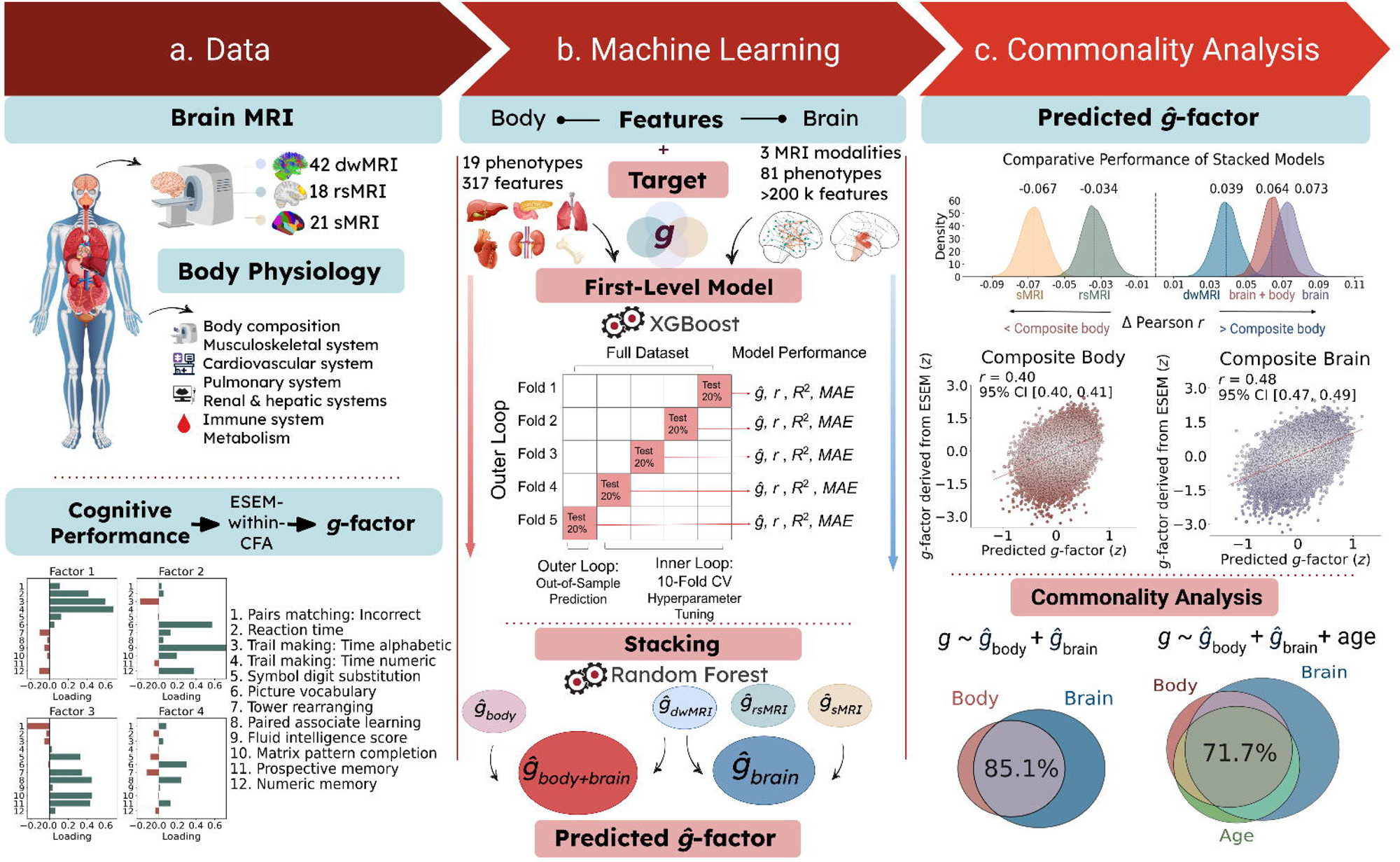
Schematic representation of the research design. **a.** Data used to derive biomarkers of cognition. Body features included nineteen phenotypes capturing body composition and seven physiological systems: cardiovascular, pulmonary, renal, hepatic, immune, metabolic, and musculoskeletal. Brain features comprised 81 neuroimaging-derived measures from three neuroimaging modalities: dwMRI, rsMRI, and sMRI. The target variable was a latent general cognition factor (*g*-factor) estimated from twelve cognitive performance scores using exploratory structural equation modelling within a confirmatory factor analysis framework (ESEM-within-CFA). **b.** Two-level machine learning framework. At the first level, we used the XGBoost algorithm to train 100 base models predicting the *g*-factor from each body and brain phenotype. At the second level, we applied multimodal stacking: *g*-factor predictions from all first-level models were used as features in Random Forest models that combined information within and across modalities. This procedure yielded six modality-specific biomarkers: a composite body marker integrating all body phenotypes (*ĝ*_body_); unimodal brain markers derived from stacking brain phenotypes within each neuroimaging modality (*ĝ*_dwMRI_, *ĝ*_rsMRI_, *ĝ*_sMRI_); a composite brain marker integrating all brain phenotypes (*ĝ*brain); and a body+brain marker combining all body and brain phenotypes (*ĝ*_body+brain_). Model performance was estimated using nested cross-validation with five outer and ten inner folds. **c.** Commonality analysis. Body- and brain-derived markers were used as explanatory variables in a series of linear regression models. The observed *g*-factor, pooled across the five outer-fold test sets, served as the response variable, while *g*-factors predicted from body, brain, or age (also pooled across outer-fold test sets) were used as explanatory variables. To quantify overlap in variance between (a) brain markers and the cognition–body relationship and (b) body and brain markers and the cognition–age relationship, we decomposed the variance of the observed *g*-factor into components explained uniquely or commonly by each pair of explanatory variables. To assess how composite body and brain markers commonly overlap with the cognition–age relationship, we extended the analysis to a model with three explanatory variables, partitioning the variance of the observed *g*-factor into components uniquely attributable to age, body, and brain, as well as variance shared by each pair of predictors and by all three predictors together. *dwMRI*, diffusion-weighted MRI; *rsMRI*, resting-state MRI; *sMRI*, structural MRI; *r*, Pearson correlation between observed and predicted *g*-factor values; *R*^2^, coefficient of determination between observed and predicted *g*-factor values; *MAE*, mean absolute error; *CV*, cross-validation; *CI*, confidence interval.

#### Predicting cognition from body phenotypes

First, we trained 19 XGBoost models to derive biomarkers of cognition from 19 body phenotypes. These biomarkers explained 0.1% to 12.4% of the variance in the *g*-factor, achieving mean Pearson correlation coefficients (*r*_mean_) ranging from 0.05 to 0.35 (Fig. 2). As indicated by bootstrapping, predictions from all body phenotypes, except heel bone densitometry, were significantly better than chance. Supplementary Table S4 lists the body phenotypes used to build the machine learning models. Supplementary Tables S5 and S6 summarize, respectively, the average cross-validated and bootstrapped predictive performance of models based on these phenotypes.

**Figure 2.**
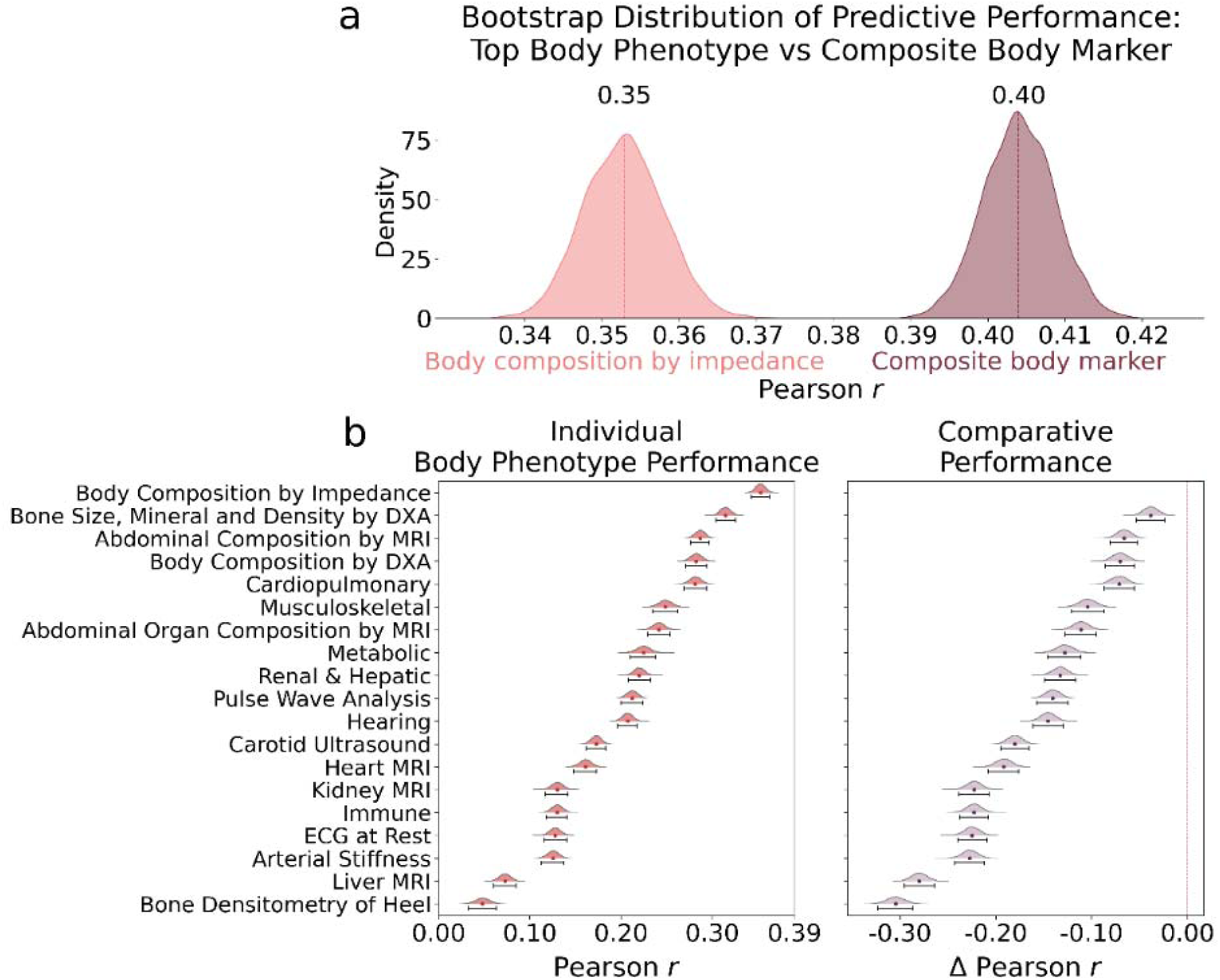
Predictive performance of each body phenotype and the composite body marker. **a.** The kernel density estimation plot shows the bootstrap distribution of predictive performance for the model based on the top-performing body phenotype (body composition by impedance) and for the model combining all body phenotypes via stacking (composite body marker). **b.** The ridgeline plots illustrate the predictive performance of each body phenotype (left) and the differences in predictive performance between each body phenotype and the top-performing phenotype (right), with corresponding 95% bootstrap confidence intervals.

Among all 19 body phenotypes, body composition assessed by impedance analysis showed the highest predictive performance, with a mean Pearson correlation of 0.35 (95% CI [0.343, 0.363]) between observed and predicted cognition. This was followed by bone mineral density (BMD) measured by dual-energy X-ray absorptiometry (DXA; *r*_mean_=0.315, 95% CI [0.304, 0.325]). In contrast, models based on liver MRI and heel bone densitometry showed the weakest predictive performance, with mean Pearson correlations of 0.073 (95% CI [0.060, 0.085]) and 0.048 (95% CI [0.033, 0.063]), respectively (Fig. 2).

Next, we employed a multimodal stacking approach to derive a composite body marker integrating all body phenotypes. This composite marker improved predictive performance, explaining 16% of cognitive variance at *r*_mean_ = 0.40 (95% CI [0.395, 0.413]) and outperforming individual phenotypes (Fig. 2). Supplementary Table S7 outlines differences in predictive performance between the composite body marker and brain-derived markers (individual brain phenotypes, unimodal brain markers, and a composite brain marker), as well as the body+brain marker.

To quantify the strength of each body phenotype’s relationship with the composite body marker, we applied a Haufe transformation (Haufe et al., 2014) and computed Pearson correlations between the composite body marker and the *g*-factors predicted from each body phenotype. Consistent with the phenotypes’ predictive performance, body composition assessed by impedance showed the strongest relevance to the composite body marker (*r* = 0.87), followed by BMD (*r* = 0.75). Liver MRI and heel bone densitometry showed the weakest associations, with Pearson *r* values of 0.19 and 0.10, respectively (Fig. 3a). Supplementary Table S8 reports the relevance of each body phenotype to the predictive performance of the stacked model and the feature importance for models based on each phenotype, computed using the Haufe transformation.

**Figure 3.**
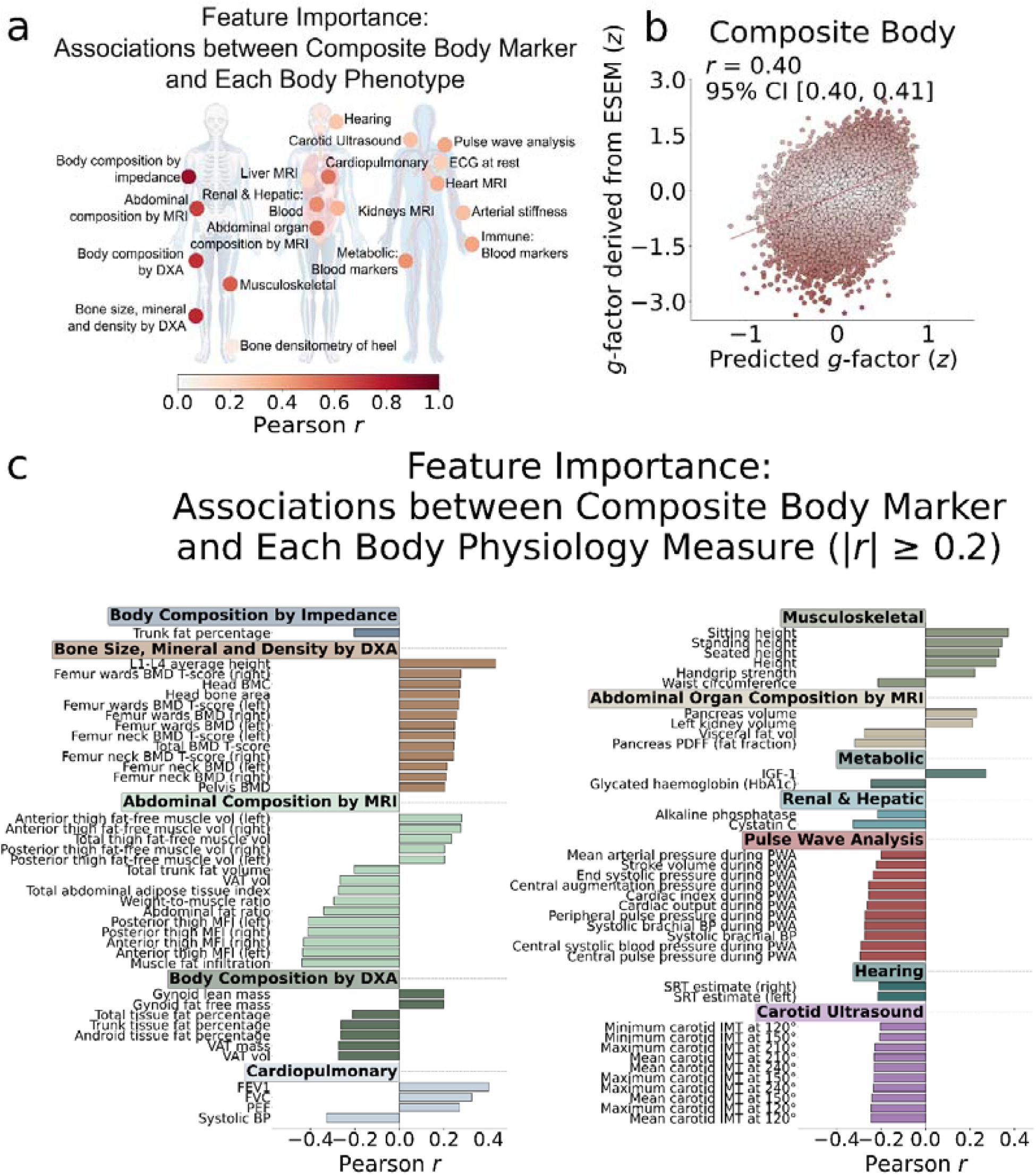
Feature importance of body phenotypes. **a.** Relationship between individual body phenotypes and the composite body marker, quantified as Pearson correlations between the *g*-factor predicted from each body phenotype and the *g*-factor predicted by the stacked model integrating all body phenotypes. **b.** Scatterplot showing the relationship between the *g*-factor derived via ESEM and the *g*-factor predicted by the stacked model integrating all body phenotypes (composite body marker). Pearson *r* reflects the mean correlation between observed and predicted *g*-factor values across the five folds, with confidence interval (CI) estimated via bootstrap resampling of the pooled observed and predicted values. **c.** Relationship between body physiology measures and the composite body marker, quantified as Pearson correlations between 317 body physiology measures and the *g*-factor predicted by the stacked model integrating all body phenotypes, pooled across the five test folds. Only correlations that remained significant after Bonferroni correction and met the threshold of *|r*| ≥ 0.2 are displayed; phenotypes not meeting this criterion are omitted. The top four correlations with |*r*| > 0.15 are reported in Table 1, and the full set of feature importance values is provided in Supplementary Table S9.

We also applied the Haufe transformation (Haufe et al., 2014) to identify which of the 317 body measures were most strongly associated with the composite body marker. The top measures for each body phenotype are summarized in Table 1 and shown in Figure 3b, and all associations are reported in Supplementary Table S9.

**Table 1.**
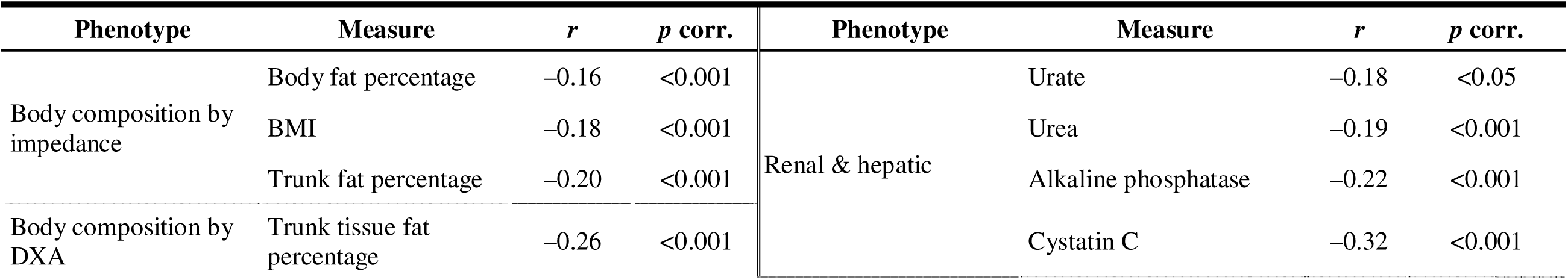
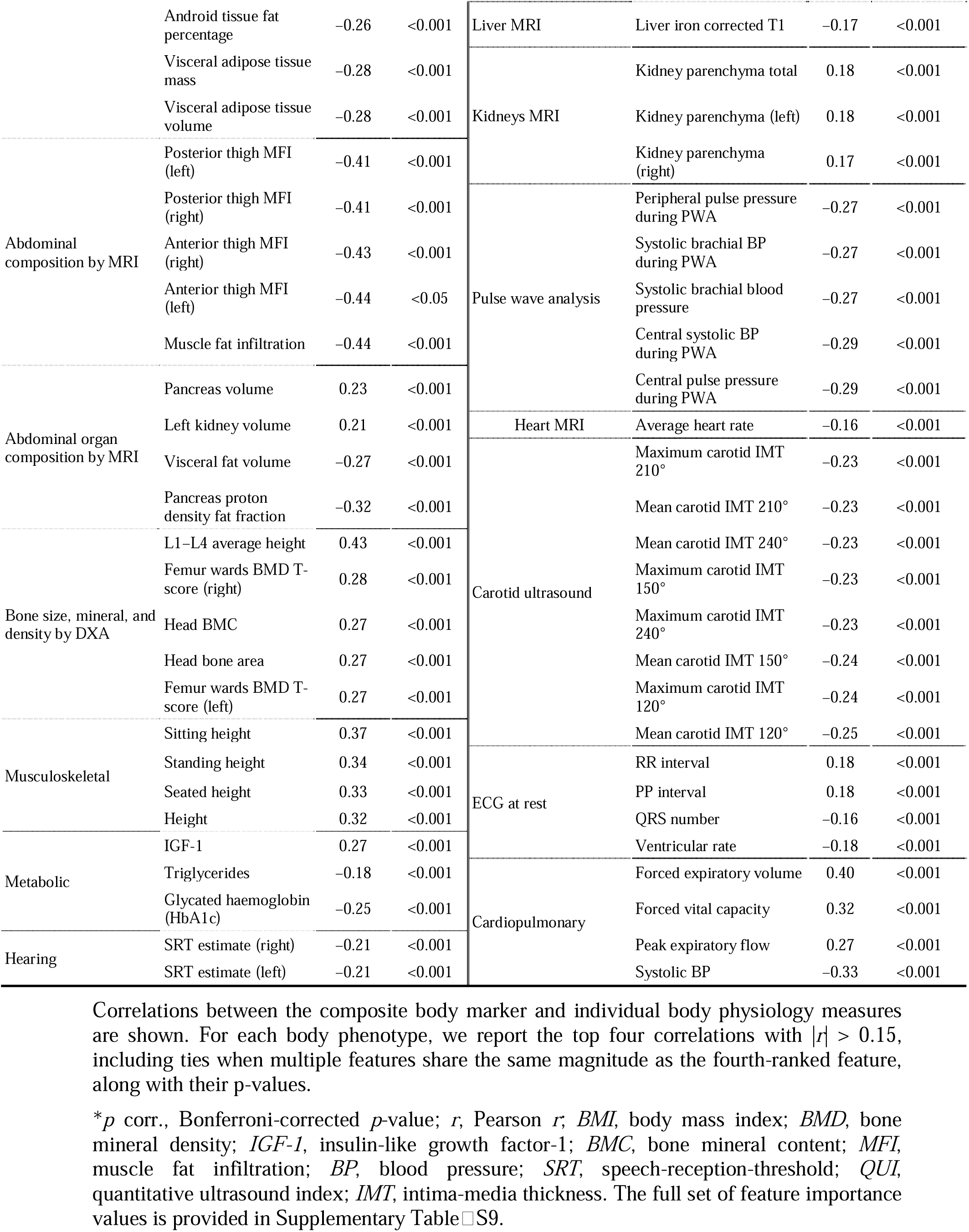
Feature importance of body physiology measures.

The strongest positive associations with the composite body marker were observed for measures of stature, bone health, and pulmonary function, whereas negative associations were found for muscle fat infiltration, ectopic fat, and blood pressure (Fig. 3b and Supplementary Table S8). For example, vertebral height in the lumbar spine (L1-L4) measured by DXA was associated with the composite marker at *r* = 0.43, followed by standing (*r* = 0.34), sitting (*r* = 0.37), and overall height (*r* = 0.32). Among measures of lung function, the strongest correlations with the composite body marker were observed for forced expiratory volume (FEV_1_; *r* = 0.40), forced vital capacity (FVC; *r* = 0.32), and peak expiratory flow (*r* = 0.27). Bone health indicators, such as femur Ward’s BMD T-score, head bone mineral content, and head bone area, showed moderate positive associations (*r* = 0.27 to 0.28), as did insulin growth factor-1 (IGF-1, *r* = 0.27).

In contrast, we observed the strongest negative associations for muscle fat infiltration in the thighs measured by MRI, with correlations ranging from *r* = −0.41 to −0.44. Moderate negative correlations with the composite body marker were also found for blood pressure and arterial load (systolic blood pressure, *r* = −0.33; central pulse pressure, *r* = −0.29), ectopic fat (pancreas fat fraction, *r* = −0.32), renal markers (cystatin C, *r* = −0.32), and adiposity measures such as weight-to-muscle ratio, visceral adipose tissue, and trunk/android fat percentages (*r* = −0.26 to −0.28). Finally, metabolic indicators, such as glycated haemoglobin (HbA1c), also showed negative associations with the composite body marker (*r* = −0.25), as did indices of vascular structure (carotid intima-media thickness, *r* = −0.25).

#### Predicting cognition from brain phenotypes

To derive brain markers of cognition, we trained machine learning models to predict the *g*-factor from 81 brain phenotypes. We then applied multimodal stacking to generate unimodal brain markers, integrating phenotypes within each of the three neuroimaging modalities, as well as a composite brain marker combining all neuroimaging data. Additionally, we built a body+brain marker by integrating all body and brain phenotypes.

Individual brain phenotypes accounted for 1.3% to 18.6% of the variance in the *g*-factor. A composite brain marker based on all brain phenotypes accounted for 23% of cognitive variance, with *r*_mean_ = 0.48 (95% CI [0.467, 0.486]). A body+brain marker integrating all body and brain phenotypes increased the explained variance to 21.8% at *r*_mean_ = 0.47 (95% CI [0.460, 0.477]). At the neuroimaging modality level, a unimodal marker based on dwMRI (*R*^2^_mean_ = 17%, *r*_mean_ = 0.44, 95% CI [0.433, 0.453]) outperformed those based on rsMRI (*R*^2^_mean_ = 9.8%, *r*_mean_ = 0.37, 95% CI [0.360, 0.381]) and sMRI (*R*^2^_mean_ = 12.8%, *r*_mean_ = 0.34, 95% CI [0.326, 0.347]) (Fig. 4).

**Figure 4.**
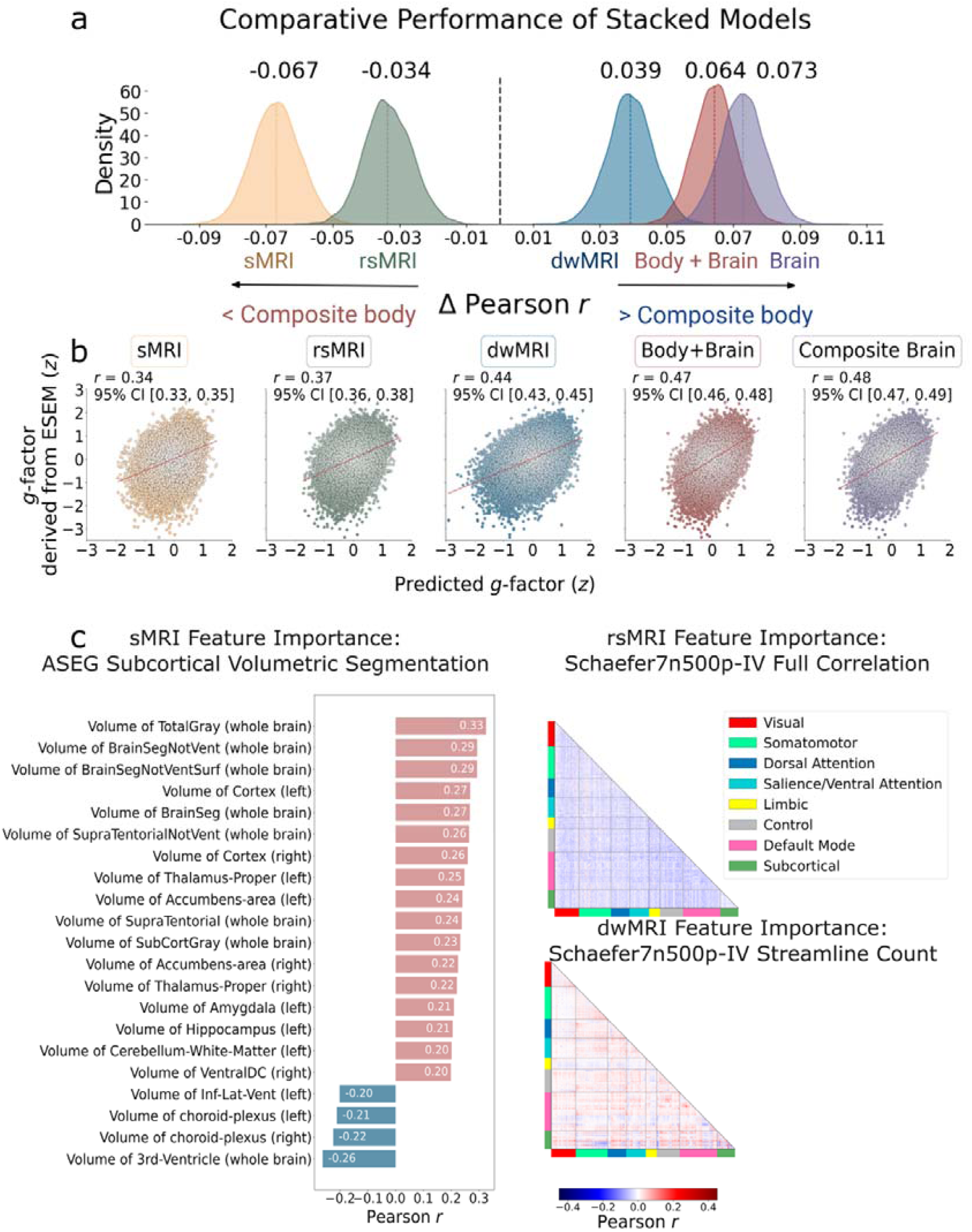
Predictive performance of body and brain markers and feature importance for top-performing brain phenotypes. **a.** Kernel density estimation plot showing the bootstrap distributions of differences (Δ) in Pearson *r* between the composite body marker and the unimodal and composite brain markers, as well as the body+brain marker. **b.** Scatterplot showing the relationship between the *g*-factor derived via ESEM and the *g*-factor predicted from brain phenotypes stacked within neuroimaging modalities (unimodal markers), across modalities (composite brain marker), and from body and brain phenotypes combined (body+brain marker). Pearson *r* reflects the mean correlation between observed and predicted *g*-factor values across the five folds, with confidence intervals (CIs) estimated via bootstrap resampling of the pooled observed and predicted values. **c.** Feature importance plots for the top-performing sMRI, rsMRI, and dwMRI phenotypes. Bar plots for sMRI show the direction and magnitude of Pearson correlations (|*r*| > 0.2) between subcortical volumes derived from FreeSurfer’s automatic subcortical segmentation (ASEG) and the composite brain marker, pooled across the five outer-fold test sets. The strongest positive associations with the composite brain marker were observed for total grey matter volume and whole-brain tissue volume excluding ventricles, cerebrospinal fluid, and dura mater. The strongest negative associations were found for the volumes of the third ventricle, choroid plexus, inferior horn of the lateral ventricle, and white matter hypointensities. Heatmaps display Pearson correlations between the composite brain marker and streamline count, as well as full correlation matrices for connections between cortical and subcortical nodes in the Schaefer7n500p-IV parcellation, pooled across the five outer-fold test sets. For dwMRI, the highest positive association with the composite brain marker was observed for structural connections between the right visual and left somatomotor areas, whereas connections between the left visual and right somatomotor regions, and between the left precuneus/posterior cingulate cortex within the default mode network (DMN) and the right visual network, showed the strongest negative associations. For rsMRI, the strongest positive association with the composite brain marker was observed for functional connectivity between the left somatomotor cortex and the right DMN parietal region. The strongest negative associations were observed for functional connectivity between the DMN left prefrontal cortex and the DMN right precuneus/posterior cingulate cortex, and between the DMN left prefrontal cortex and the right lateral prefrontal cortex of the control network. Full results of the feature importance analysis for the top-performing brain phenotypes are summarized in Supplementary Tables S12–S14.

Among brain phenotypes from dwMRI and rsMRI, the number of streamlines and functional connectivity among cortical and subcortical regions within the Schaefer7n500p-IV parcellation showed the highest predictive performance for the *g*-factor, with *r*_mean_ = 0.43 (95% CI [0.421, 0.441]) and *r*_mean_ = 0.34 (95% CI [0.326, 0.348]), respectively, explaining 18.6% and 11% of the variance. For sMRI, subcortical volumes derived from FreeSurfer’s automatic subcortical segmentation (ASEG) outperformed other structural phenotypes, predicting cognition at *r*_mean_ = 0.32 (95% CI [0.306, 0.327]) and explaining 10% of cognitive variance (Fig. 4). Supplementary Tables S10 and S11 present the average cross-validated performance and the bootstrapped predictive performance, respectively, for models derived from individual brain phenotypes and for the stacked brain models. The results of the feature importance analysis for the top-performing brain phenotypes are summarized in Supplementary Tables S12–S14.

We benchmarked the predictive performance of the composite body marker against unimodal brain markers, a composite brain marker, and a body+brain marker by comparing their bootstrapped performance estimates. The composite brain marker (*Δr* = 0.073, 95% CI [0.06, 0.086]), body+brain marker (*Δr* = 0.064, 95% CI [0.052, 0.076]), and dwMRI-based marker (*Δr* = 0.039, 95% CI [0.026, 0.052]) all outperformed the composite body marker in predicting cognition. In contrast, models based on rsMRI (*Δr* = −0.033, 95% CI [−0.047, −0.019]) and sMRI (*Δr* = −0.067, 95% CI [−0.081, −0.053]) showed inferior performance relative to body alone (Fig. 4).

### Commonality Analyses

#### Cognition–body relationship overlapping with brain markers

We conducted commonality analyses to quantify the proportion of the cognition–body covariance that overlapped with brain markers. For unimodal brain markers (phenotypes stacked within each neuroimaging modality) and the composite brain marker (phenotypes stacked across all modalities), this proportion was calculated as the overlapped variance in the observed *g*-factor, commonly explained by both body and brain markers, divided by the total variance in the observed *g*-factor explained by body markers. The results of the commonality analyses for individual body phenotypes and a composite body marker are presented in Supplementary Table S15. Supplementary Table S16 further reports the corresponding analyses stratified by sex.

The cognition–body covariance substantially overlapped with brain markers. A composite brain marker integrating all brain phenotypes across three neuroimaging modalities captured 85.1% of the covariation between a composite body marker and cognition. Among the three neuroimaging modalities, dwMRI captured the greatest portion of cognition–body covariance, accounting for 86.2% of the relationship. In contrast, sMRI (57.2%) and rsMRI (49.5%) accounted for about half of the cognition–body covariance (Fig. 5).

**Figure 5.**
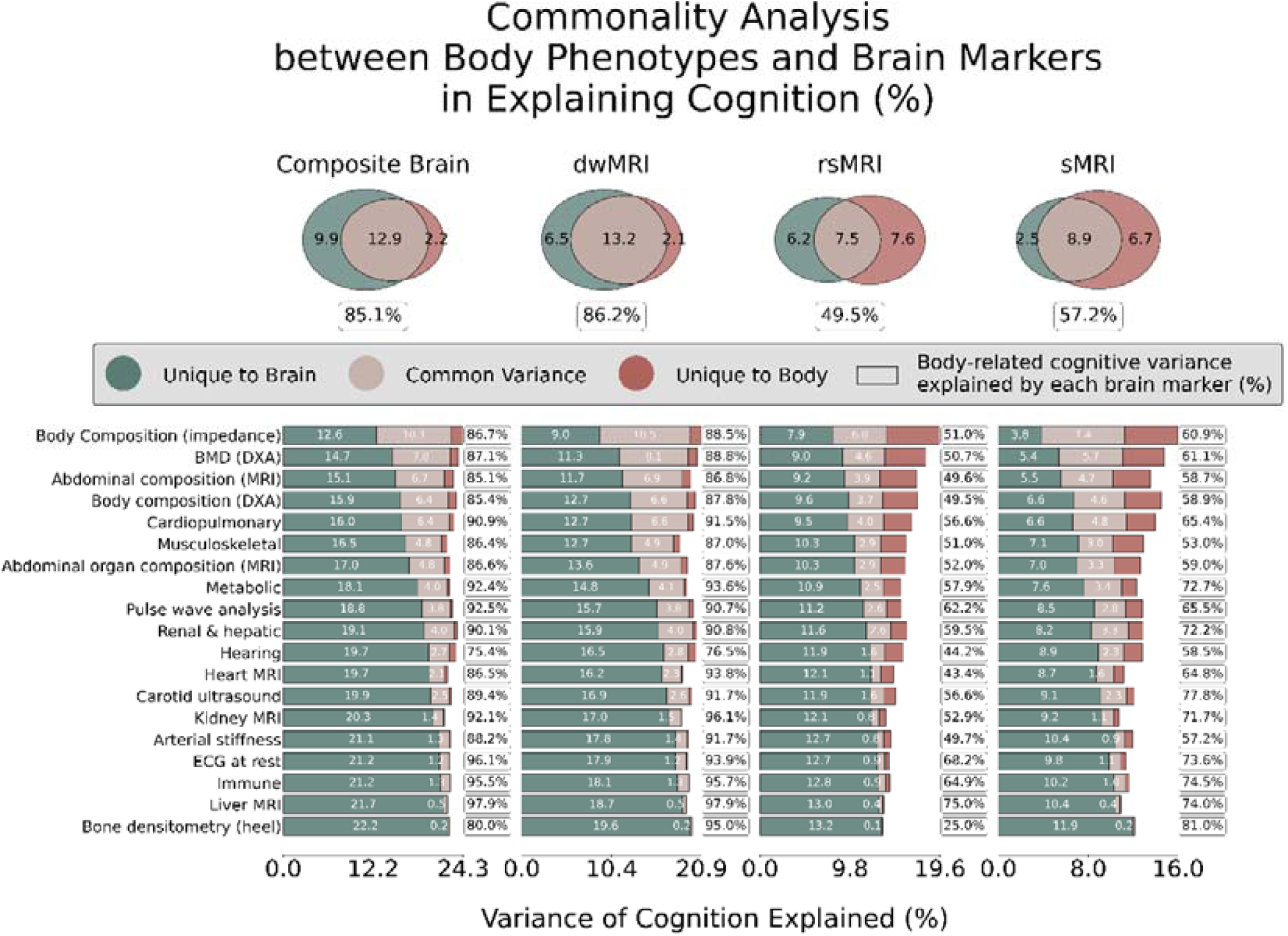
Commonality analysis between the body and brain in explaining cognition. Venn diagrams (top) illustrate the unique and common variance between the composite body marker and brain markers in explaining cognition (observed *g*-factor). Percentages in the box indicate the proportion of cognition–body covariance overlapping with each brain marker, calculated as: ***% Brain* = *Common_composite body, composite brain_* ÷ (*Common_composite body, composite brain_ + Unique_body_*)** Stacked bar plots (bottom) represent the unique and common variance between individual body phenotypes and brain markers in explaining cognition (observed *g*-factor). Percentages in the box indicate the proportion of covariation between each body phenotype and cognition that is explained by a given brain marker. *Unique variance* refers to the proportion (%) of variance in the *g*-factor explained exclusively by body or brain, whereas *common variance* reflects the proportion (%) of variance in the *g*-factor jointly explained by both.

The cognition–body covariance showed the greatest overlap with dwMRI phenotypes, accounting for over 75% of the covariation. For example, dwMRI phenotypes captured more than 90% of the cognitive variance associated with liver, kidney, and cardiopulmonary functions, cardiovascular health, and blood markers of immune and metabolic status, and over 85% of the variance attributed to body composition and BMD in the spine and peripheral skeleton.

In comparison, sMRI phenotypes overlapped with cognition–body covariance to a lesser extent (53–81%), and most rsMRI phenotypes showed overlap below 65%. Notable exceptions for rsMRI included liver MRI and resting ECG, which demonstrated higher overlaps of 75% and 68%, respectively.

For certain body phenotypes, such as liver MRI and heel bone densitometry, the composite brain markers accounted for over 95% of the covariance with cognition despite minimal common variance. This pattern likely reflects the even smaller unique variance of these body measures relative to their common variance (Fig. 5).

#### Cognition–age relationship overlapping with body physiology and brain markers

We employed commonality analysis to quantify the proportion of the cognition–age covariance that overlapped with body and brain markers. The analysis showed that the cognition–age covariance substantially overlapped with the composite body (81.4%) and brain (87.2%) markers. The common variance explained jointly by these composite body and brain makers was 71.7%. Together, the composite body and brain markers explained 96.8% of the cognition–age covariance (Fig. 6 and Supplementary Table S17).

**Figure 6.**
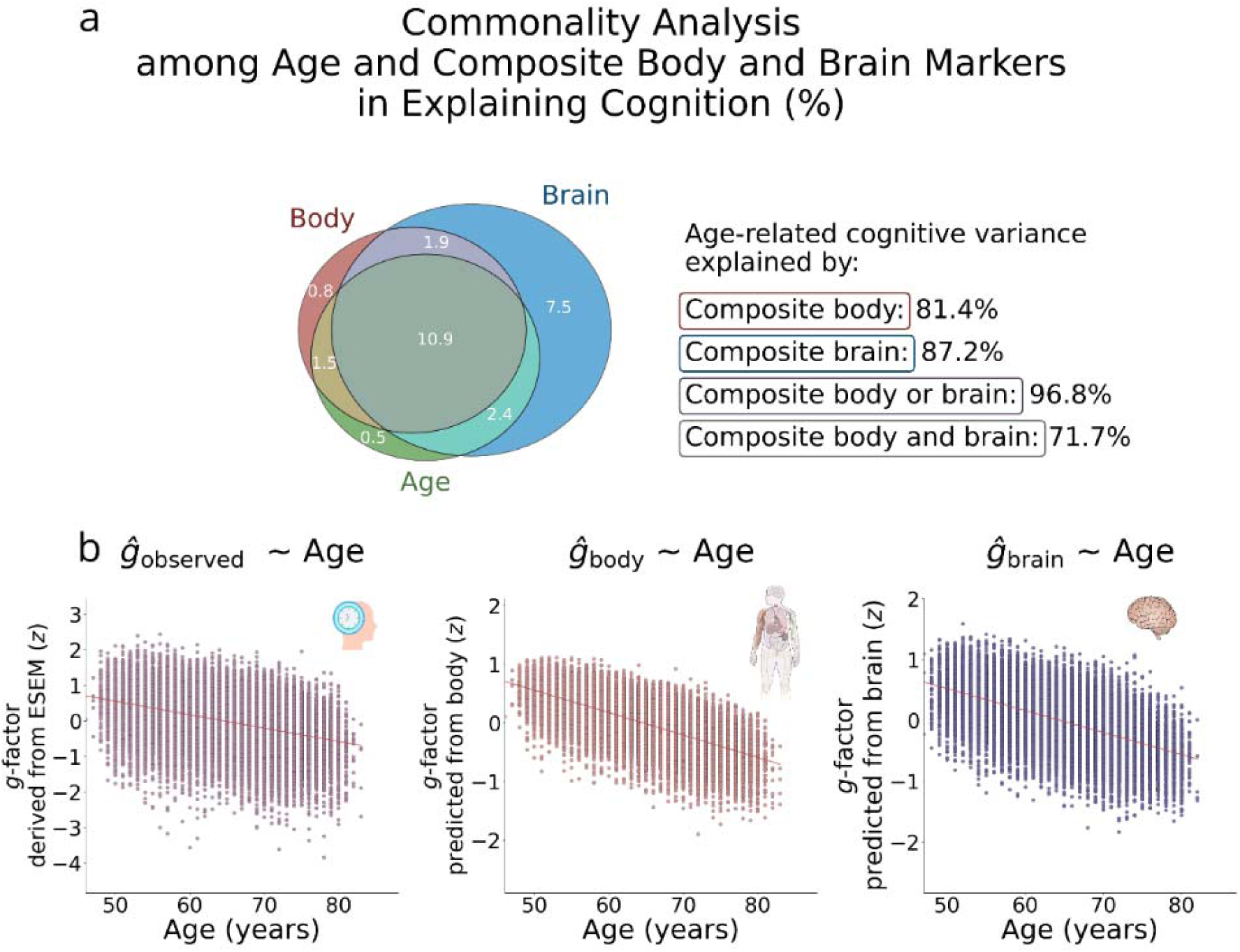
Commonality analysis among age and composite body and brain markers in explaining cognition. **a.** Venn diagram showing the unique and shared contributions of age, body, and brain to cognitive variance. The proportion of age-related cognitive variance explained by body and brain was computed as follows: **% Body** = (Common_age,body_ + Common_age,body,brain_) **÷** (Unique_age_ + Common_age,body_ + Common_age,body,brain_ + Common_age,brain_) **% Brain** = (Common_age,brain_ + Common_age,body,brain_) **÷** (Unique_age_ + Common_age,body_ + Common_age,body,brain_ + Common_age,brain_) **% Brain or body** = (Common_age,body_ + Common_age,brain_ + Common_age,body,brain_) **÷** (Unique_age_ + Common_age,body_ + Common_age,body,brain_ + Common_age,brain_) **% Brain and body** = (Common_age,body,brain_) **÷** (Unique_age_ + Common_age,body_ + Common_age,body,brain_ + Common_age,brain_) **b.** Scatterplots showing the *g*-factor derived via ESEM (*r_age_*= –0.40, *R*^2^*_age_* = 0.16, *p* = <0.001) and the *g*-factor predicted from the stacked body phenotypes (composite body; *r_age_* = –0.70, *R*^2^*_age_* = 0.49, *p* = <0.001) and stacked brain phenotypes (composite brain; *r_age_* = –0.58, *R*^2^ *_age_* = 0.34, *p* = <0.001), each plotted as a function of age.

Among individual body and brain phenotypes, cognition–age covariance showed the greatest overlap with structural connectome measures derived from dwMRI, exceeding 69%. This was followed by structural properties of subcortical regions from FreeSurfer volumetric segmentation (ASEG volume and mean intensity) and regional grey matter volumes from FSL FAST, which accounted for 53.2–60.7% of the covariance. Notably, body composition measured via impedance analysis showed comparably high overlap (68.5%), outperforming sMRI phenotypes, while BMD assessed using DXA exhibited a 51.5% overlap.

In contrast, several measures, including partial correlations of BOLD signals from the Schaefer7n500p-IV parcellation, amplitudes of 21 resting-state components, diffusion tensor mode from probabilistic tractography, cortical surface areas from FreeSurfer *ex-vivo* Brodmann parcellation, and a range of body physiology indices (e.g., kidney and liver MRI, immune markers, resting ECG, arterial stiffness, and heel bone densitometry), each overlapped by less than 10% with the cognition–age covariance (Fig. 7). Results are summarized in Supplementary Table S18, with sex-stratified analyses provided in Supplementary Table S19.

**Figure 7.**
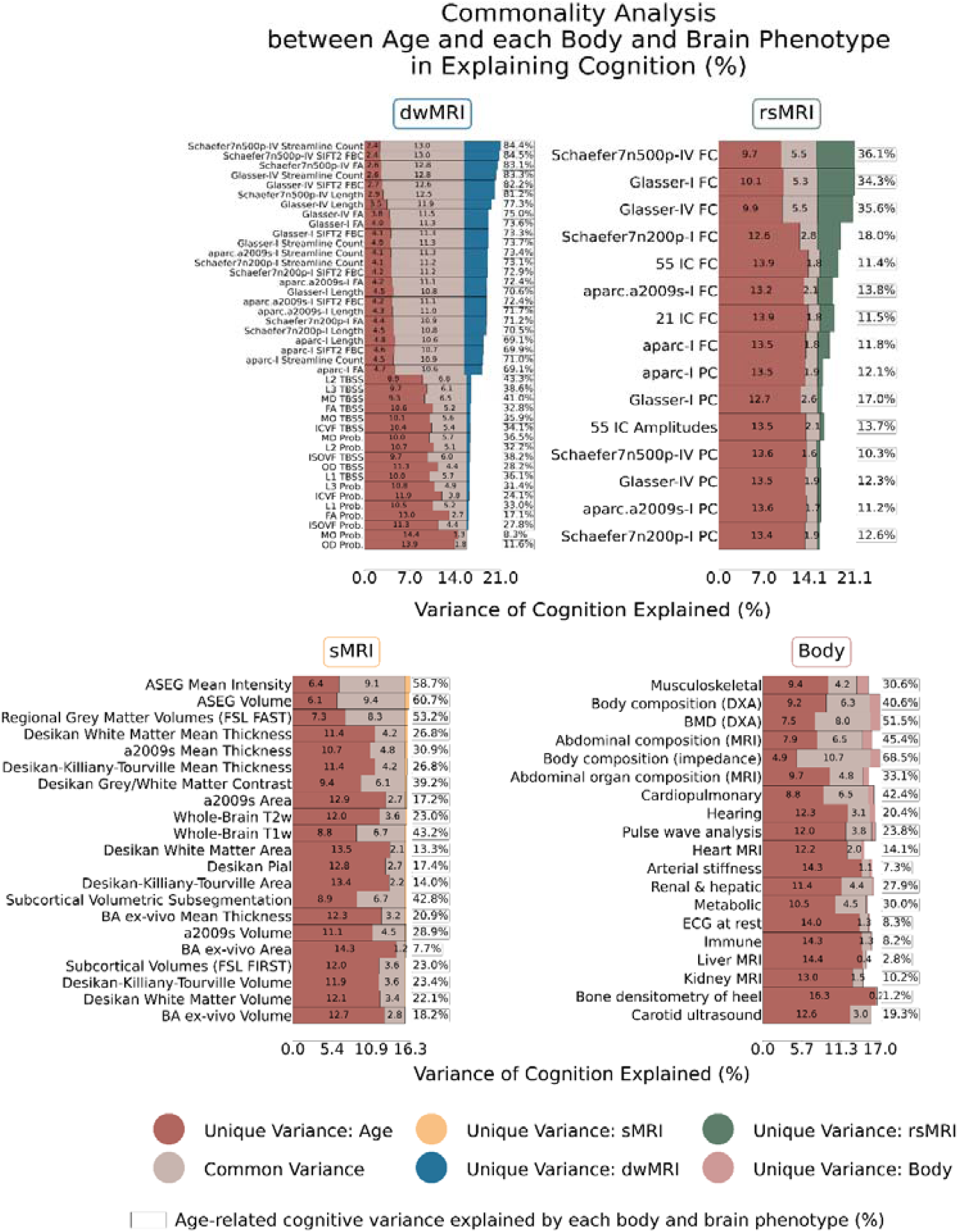
Commonality analysis between age and each body and brain phenotype in explaining cognition. Stacked bar plots represent the unique and common variance between age, individual body phenotypes, and individual brain phenotypes in explaining cognition (observed *g*-factor). Percentages in the box indicate the proportion of covariation between age and cognition explained by each body and brain phenotype. *Unique variance* refers to the proportion (%) of variance in the *g*-factor explained exclusively by age, body, or brain, whereas *common variance* reflects the proportion (%) of variance in the *g*-factor jointly explained by age, body, and brain. *Schaefer7n500p*, Schaefer cortical atlas – 7 networks, 500 parcels; *Schaefer7n200p*, Schaefer cortical atlas – 7 networks, 200 parcels; *aparc.a2009s*, Destrieux cortical atlas; *I*, Melbourne Subcortical Atlas I; *IV*, Melbourne Subcortical Atlas IV; *SIFT2*, Spherical-Deconvolution Informed Filtering of Tractograms 2; *Prob.*, probabilistic; *TBSS*, Tract-Based Spatial Statistics; *FA*, fractional anisotropy; *MD*, mean diffusivity; *MO*, diffusion tensor mode; *L1*, *L2*, *L3*, eigenvalues of the diffusion tensor; *OD*, orientation dispersion index; *ICVF*, intracellular volume fraction; *ISOVF*, isotropic volume fraction; *IC*, independent components; *FC*, full correlation; *PC*, partial correlation; *ASEG*, FreeSurfer automated subcortical volumetric segmentation; *FSL FAST*, FMRIB’s Automated Segmentation Tool; a2009s, Destrieux atlas; *T2w*, T2-weighted MRI; *T1w*, T1-weighted MRI; *BA*, FreeSurfer *ex-vivo* Brodmann Area Maps; *FSL FIRST*, FMRIB’s Integrated Registration and Segmentation Tool; *DXA*, dual-energy X-ray absorptiometry; *BMD*, bone mineral density; *MRI*, magnetic resonance imaging; *ECG*, electrocardiogram.

## DISCUSSION

Our study had three primary aims. First, we evaluated the predictive performance of body phenotypes for cognition. Using a composite body marker that integrated 317 measures across 19 phenotypes, we predicted cognitive functioning with a correlation of *r* = 0.40, demonstrating a cognition–body covariance at 16%. Combining body phenotypes with brain neuroimaging enhanced the predictive power to *r* = 0.47. Second, we quantified the extent to which the cognition–body covariance overlapped with neuroimaging measures. We found that 85.1% of the cognition–body covariance was shared with a composite brain marker aggregating 81 neuroimaging phenotypes. Third, to extend the analysis to cognitive aging, we examined how the cognition–age relationship overlapped with body physiology and neuroimaging measures. The cognition–age covariance showed 71.7% joint overlap with the composite brain and body markers, and, when considered together, these markers accounted for 96.8% of the cognition–age covariance.

### Predictive performance of body phenotypes

A composite body marker integrating 19 body phenotypes allowed us to quantify the overall strength of the cognition–body relationship, explaining 16% of variance in cognition. Its predictive performance (*r* = 0.40) exceeded that of rsMRI and sMRI but was lower than that of dwMRI and the composite brain marker, which captured 23% of variance in cognition (*r* = 0.48). These effect sizes are consistent with previous literature on brain–cognition associations (Dhamala et al., 2021; Krämer et al., 2024; Pat et al., 2022; Tetereva et al., 2022), whereas studies examining predictive body–cognition relationships remain comparatively limited. Although a substantial proportion of cognitive variance remained unexplained, all reported associations were assessed out-of-sample, that is, in individuals not included in model training, where predictive performance and explained variance are typically attenuated relative to in-sample estimates. Taken together, these findings position body physiology as a promising predictive marker of cognitive functioning and provide quantitative evidence for its association with cognition (Marek et al., 2022).

By examining the predictive performance of individual body physiology phenotypes and the feature importance of their stacked model, we benchmarked the relevance of each phenotype to cognitive health. While the performance varied widely, predictions from 18 of the 19 body physiology phenotypes were significantly better than chance, consistent with studies linking cognitive functioning to body physiology when each phenotype is examined in isolation (Gao et al., 2024; Koutsonida et al., 2022; Ma et al., 2025; Zou et al., 2025). This confirms the roles of body composition (Chen et al., 2024), cardiovascular (Song et al., 2020), pulmonary (Duggan et al., 2020), renal (Madero et al., 2008), hepatic (Kjærgaard et al., 2021; Zhang et al., 2024), immune (Cunha et al., 2026; Liu et al., 2024), metabolic (Dimache et al., 2021; Koutsonida et al., 2022), and musculoskeletal (Laudisio et al., 2016) systems in cognition, while also quantifying the strength of their out-of-sample relationship with cognitive functioning.

Among the 19 body phenotypes examined, body and abdominal composition, as well as the musculoskeletal system, were the strongest predictors of cognition, rivalling brain markers derived from rsMRI and sMRI. The predictive power of these phenotypes was mainly driven by body adiposity and bone size, mineral content, and density. For example, body and trunk fat percentage, muscle fat infiltration, body mass index (BMI), and visceral adipose tissue (VAT) volume and mass were negatively associated with the composite body marker, whereas lean and fat-free mass, height, as well as L1–L4 average height and femur and head BMD showed positive associations. These features have been consistently associated with cognitive and brain health in later life (Chen et al., 2024; Laudisio et al., 2016; Samuelsson et al., 2025; Zhang et al., 2025) and are suggested to reflect common pathways linking systemic physiology and brain integrity (Cole, 2020; Cole et al., 2018, 2019; Samuelsson et al., 2025). For example, bone mass loss is considered a risk factor for white matter changes, suggesting shared biological pathways underlying brain and bone health (Minn et al., 2014). This assumption is further supported by the positive correlation between height and predicted cognition. Although this association may arise from multiple influences (Aguiar-Oliveira et al., 2026; Amin & Fletcher, 2022), prior research shows that the height–cognition relationship remains robust even after accounting for genetic, socioeconomic, and health-related factors, pointing to cumulative childhood nutrition as a key driver (Amin & Fletcher, 2022; Kobayashi et al., 2019).

The negative associations we observed for body and trunk fat percentage, muscle fat infiltration, BMI, and VAT volume are consistent with evidence linking adiposity to poorer brain health and cognitive outcomes (Rosano et al., 2023; Sui & Pasco, 2020; Tian et al., 2024). For instance, prior studies have reported relationships between regional fat accumulation and reduced grey matter volume, white matter lesions, and amyloid-*β* burden (Anand et al., 2022; Beller et al., 2019; Boccara et al., 2023; Kim et al., 2022; McEwen, 2002). The underlying mechanisms are widely discussed in the literature (Wu et al., 2025) and may involve excessive leptin secretion impairing amyloid precursor protein transport (Fewlass et al., 2004), allostatic load (McEwen, 2002), systemic inflammation (Cannavale et al., 2021; Tian et al., 2024), and metabolic dysregulation (Gu et al., 2025). In contrast, lean and fat-free muscle mass, showing positive associations with predicted cognition, may exert neuroprotective effects (Wu et al., 2025) through improved glucose and lipid metabolism and the release of myokines that regulate neurotrophins (Oudbier et al., 2022; Taha et al., 2022; Tian et al., 2024).

Abdominal organ composition, cardiopulmonary function, kidney and liver health, and metabolic markers likewise showed high relevance to the composite body marker. At the feature level, lung capacity (FEV and FVC) and circulating IGF-1 showed the strongest positive associations, whereas ectopic fat deposition (including pancreas and liver fat), elevated blood pressure, impaired kidney function, and dysregulated glucose and lipid metabolism were linked to poorer predicted cognition. Metabolic markers, such as IGF-1 and HbA1c, may be a potential bridge linking body composition with brain and cognitive function (Aguiar-Oliveira et al., 2026). IGF-1 supports neurogenesis, oligodendrocyte growth, and myelination, and may also indirectly influence brain integrity by regulating cerebral blood flow, glucose metabolism, and vascular remodelling (Aguiar-Oliveira et al., 2026). For example, IGF-1 levels exhibit a U-shaped relationship with risks of dementia and stroke, alongside a positive linear association with Parkinson’s disease (Cao et al., 2023). Notably, IGF-1 levels have been shown to exhibit non-linear associations with age (Picca et al., 2025). Similarly, increased HbA1c, crucial for glycaemic control, has been linked to grey matter abnormalities and cognitive impairment, particularly in older adults (Xiao et al., 2025; Xu et al., 2023; Zheng et al., 2023). Potential mechanisms discussed in the literature include amyloid-*β* accumulation, *tau* hyperphosphorylation, neuroinflammation, oxidative stress, immune activation, vascular dysfunction, and impaired neuronal energy metabolism, which may collectively contribute to vascular complications, brain injury, and ultimately neurodegeneration and cognitive impairment (Kellar & Craft, 2020, 2020; Xiao et al., 2025; Zheng et al., 2023).

Numerous studies have linked lung function to cognitive ability (Grande et al., 2024; Hu et al., 2025; Russ et al., 2020; Wang et al., 2022). Although the underlying mechanisms remain debated, impaired pulmonary function may compromise brain and cognitive health through hypoxia, vascular lesions, immune dysregulation, or reflect shared lifestyle factors and concurrent pathologies such as cerebrovascular disease (Huang et al., 2025; Morgan et al., 2018; Zhang et al., 2025). Liver and pancreas disorders have likewise been associated with cognitive decline and increased risk of Alzheimer’s dementia, potentially through neuroinflammation, oxidative stress, endothelial dysfunction, insulin resistance, vascular white matter lesions, and reduced brain volume (Filipović et al., 2018; Golan Shekhtman et al., 2024; Muthulingam et al., 2022; Petta et al., 2016; Sun et al., 2023). Another factor that may contribute to vascular brain lesions is hypertension. Chronic high blood pressure induces structural remodelling of cerebral vessels, leading to small vessel disease, impaired cerebral perfusion, white matter damage, lacunes, and microinfarcts – pathological changes that are associated with increased *tau* burden and vascular cognitive impairment (Cooper et al., 2022; Hu et al., 2022; Pacholko & Iadecola, 2024). These alterations may disrupt neurovascular coupling and blood–brain barrier integrity, thereby increasing vulnerability to cognitive decline in aging populations. Together, these findings underscore the importance of developing tailored approaches that incorporate system-wide physical health indicators that shape cognitive outcomes in later life.

### How much of the cognition–body relationship overlaps with brain phenotypes

Using commonality analyses, we assessed how similarly brain and body measures relate to cognition. Of the total cognition–body covariance (*r* = 0.40, *R*^2^ = 0.16), 85.1% overlapped with composite brain markers derived via multimodal stacking, highlighting the shared contribution of brain measures across physiological systems. Even for systems with relatively low standalone predictive power, the majority of their explained variance was shared with brain phenotypes. For example, the renal and hepatic systems accounted for only 4.5% of the variance in cognition independently, yet most of this variance (90.1%) was shared with the brain measures.

Among the three neuroimaging modalities, dwMRI showed the strongest overlap with the cognition–body relationship at 86.2% of the covariation. These findings support a critical role of white matter in enabling communication between brain regions, which is essential for cognitive functioning (Filley & Fields, 2016). Notably, alterations in white matter have been identified as stronger predictors of cognitive aging than changes in grey matter and may even precede grey matter loss (Celle et al., 2022; Hong et al., 2016). Furthermore, numerous studies have reported associations between white matter pathology and body health, including cardiometabolic and cardiopulmonary function (Grosu et al., 2021; Ho et al., 2018; Scharf et al., 2019). Our results reinforce these findings by identifying white matter characteristics as key neurobiological correlates of the cognition–body relationship.

Within dwMRI, streamline counts from the Schaefer7n500p-IV parcellation showed the highest predictive performance for cognition, with both positive and negative contributions from pathways linking visual, somatomotor, and default mode network (DMN) regions. In particular, the most prominent positive association with the composite brain marker was observed for the structural connections between the right visual and left somatomotor areas, whereas connections between the left visual and right somatomotor regions, and between the left precuneus/posterior cingulate cortex (PCC) within the default mode network (DMN) and the right visual network, showed negative associations. These regions play a crucial role in supporting cognitive abilities. The posterior cingulate cortex is involved in regulating internally directed cognition, attention focus, awareness, and change detection (Leech & Sharp, 2014), which also requires connections to visual areas. Likewise, the precuneus is associated with visuospatial and self-referential processing, episodic memory retrieval, and consciousness (Jitsuishi & Yamaguchi, 2023). Somatomotor areas participate in sensorimotor integration and motor control, and their strong reciprocal connections with visual areas, essential to perception and attention (Cook et al., 2025), form key pathways supporting cognitive functioning (Dresp-Langley, 2022; El Rassi et al., 2024).

It is important to note that a substantial portion of the cognition–body relationship also overlapped, although not as high, with rsMRI (49.5%) and sMRI (57.2%). This suggests the important roles of functional connectivity and grey matter morphology. rsMRI is widely used to predict individual differences in cognitive abilities (Chen et al., 2022; Chén et al., 2019; Joshi et al., 2019; Kong et al., 2019; Sripada et al., 2020; Wehrheim et al., 2023) and is considered a valuable tool for studying cognitive impairments (Li et al., 2018). Resting-state brain activity reflects multiple cognitive processes, including attention (Machner et al., 2022) and memory (Osaka et al., 2021).

Within rsMRI, functional connectivity between the somatomotor and DMN regions, as well as bilateral connectivity within the DMN, showed the strongest associations with the composite brain marker. Specifically, connectivity between the left somatomotor cortex and the right DMN parietal region was positively associated with the marker, whereas connectivity of the left prefrontal cortex of the DMN and the right precuneus/PCC DMN node, as well as the right lateral prefrontal cortex of the control network, showed the strongest negative associations. These networks and regions are central to cognitive processing: the lateral prefrontal cortex is involved in goal-directed and stimulus-driven attention and memory (Asplund et al., 2010; Blumenfeld & Ranganath, 2019), and the parietal cortex is pivotal for spatial attention, sensory integration, and number processing (Bisbing et al., 2015; Brass et al., 2005; Katsuki & Constantinidis, 2012).

Resting-state brain activity is closely intertwined with body physiology. Autonomic arousal and slow cardiorespiratory fluctuations influence both local neural activity and large-scale functional connectivity, whereas cardiovascular and metabolic changes, as well as aging, contribute to alterations in network organization and physiological rsMRI signatures. For example, elevated heart rate variability has been associated with increased functional connectivity, particularly stronger coupling of the brainstem, thalamus, putamen, and dorsolateral prefrontal cortex with the amygdala and dorsal anterior cingulate cortex, suggesting distinct contributions of parasympathetic and sympathetic tone (Chang et al., 2013) (Chang et al., 2013). Cardiovascular and metabolic health have also been linked to whole-brain functional network integrity. In the Human Connectome Project Aging study, poorer cardiometabolic health scores were associated with reduced global functional connectivity, with especially pronounced effects in insular, medial frontal, medial parietal, and superior temporal regions. Cardiometabolic risk was particularly strongly linked to altered connectivity between the DMN, cingulo-opercular, and dorsal attention networks (Rashid et al., 2023). Moreover, metabolic syndrome has been associated with disrupted intrinsic communication within core neural networks and altered between-network connectivity across the brain, including hyperconnectivity and reduced network modularity, potentially reflecting the cumulative impact of co-occurring vascular risk factors (Rashid et al., 2021). Mendelian randomization analyses in the UK Biobank further indicate bidirectional causal links between specific resting-state networks and cardiovascular diseases, including hypertension, atrial fibrillation, heart failure, and coronary artery disease (Yang et al., 2025). Recent work further suggests that vascular regulation may contribute directly to resting-state functional connectivity, as vascular fluctuations linked to respiration and CO_2_ dynamics can spatially mirror canonical functional brain networks, including the DMN, task-positive, and visual networks (Bright et al., 2020).

Finally, within sMRI, the ASEG parcellation showed the greatest predictive performance for cognition. Volumes of total grey matter and whole brain tissue excluding ventricles, cerebrospinal fluid, and dura mater were positively associated with the composite brain marker, whereas volumes of the third ventricle, choroid plexus, inferior horn of the lateral ventricle, and white matter hypointensities showed negative associations. These findings align with prior research demonstrating positive relationships between grey matter and total brain volumes and cognitive abilities (de Chastelaine et al., 2023; Royle et al., 2013; Taki et al., 2011). In contrast, ventricular enlargement, particularly of the third ventricle, has been identified as a risk factor for cognitive impairment (Gao et al., 2024; Lee et al., 2024). Increased choroid plexus volume, a vascularised tissue involved in cerebrospinal fluid production, has likewise been linked to a range of brain disorders and cognitive decline (Diez-Cirarda et al., 2025; Jeong et al., 2023; Tireli et al., 2025). White matter hypointensities, reflecting white matter lesions, are also well-established correlates of cognitive impairment and abnormal amyloid-*β* (Hedberg et al., 2023; Wei et al., 2019).

Structural MRI studies also demonstrate close links between brain morphology and body physiology, particularly body adiposity, muscle mass, and metabolic function. Such associations are reflected in variations in cortical thickness, surface area, and regional brain volumes. For example, in the UK Biobank cohort, greater body weight and adiposity were associated with widespread morphological alterations, including nonlinear reductions in cortical and cerebellar grey matter volumes, smaller brainstem volumes, larger ventricular volumes, and larger accumbens volume, whereas greater thigh muscle volume was associated with better structural measures (Gurholt et al., 2021). Regionally, higher body mass index has been linked to focal cortical thinning, particularly in ventromedial prefrontal and lateral occipital regions (Medic et al., 2016). Beyond anthropometric measures, body MRI-derived indices of adiposity, such as liver fat and muscle fat infiltration, have been associated with lower cortical area, thickness, and volume, as well as reduced cerebellar volume, while muscle volume has shown the opposite pattern, relating to more preserved brain tissue (Gurholt et al., 2021). Cardiometabolic risk factors, including diabetes, hypertension, and hypercholesterolemia, have similarly been linked to less favorable structural brain profiles, suggesting that long-term metabolic and vascular health may contribute to widespread variation in brain morphology (Gurholt et al., 2021, 2024). Although the underlying mechanisms remain incompletely understood, vascular and metabolic processes, including altered cerebrovascular health, systemic inflammation, insulin resistance, and dysregulated energy metabolism, may contribute to associations between body physiology and brain anatomy and morphology (Coucha et al., 2018; Vinuesa et al., 2021).

Collectively, these findings advance our understanding of how different brain characteristics shape the relationship between body and cognition, and they open avenues for future work to identify strategies that enhance brain resilience to age-related factors such as adiposity, metabolic dysregulation, and muscle loss.

### How much of the cognition–age relationship overlaps with body and brain phenotypes

Using a separate set of commonality analyses, we examined the extent to which cognition–age covariance was shared with composite body and brain markers. Of the total cognition–age covariance (*r* = −0.40, *R*^2^ = 0.16), 81.4% and 87.2% overlapped with the composite brain and body markers, respectively. When considered together, nearly all cognition–age covariance overlapped with either brain or body markers (96.8%). The remaining unexplained variance may reflect the inherent noisiness of body physiology, neuroimaging, and cognitive measures (Elliott et al., 2020; Enkavi et al., 2019; Hermes et al., 2017).

Notably, most of the explained variance was shared between the composite body and brain markers (71.7%), indicating that body and brain phenotypes capture highly overlapping aspects of age-related cognitive differences. Taken together, these findings suggest that age-related cognitive variation is strongly associated with shared patterns spanning systemic physiology and the brain. These insights suggest potential translational relevance. They provide a holistic understanding of the biology of individual variations in cognitive functioning in older adults. Preventive interventions, ranging from cardiometabolic monitoring and pulmonary rehabilitation to nutritional support and promotion of physical activity, may reduce the burden of cognitive decline if implemented early and sustained across the lifespan. Moreover, multimodal biomarker frameworks that combine systemic physiology with neuroimaging could improve risk stratification, guide personalised interventions, and inform clinical trials aimed at delaying or mitigating cognitive aging.

More broadly, these findings support geroscience perspectives suggesting that cognitive aging involves coordinated changes across multiple biological pathways. Previous research shows that blood-based metabolic, inflammatory, vascular, and neurodegenerative markers follow distinct age-related trajectories over time, with inflammatory and metabolic systems showing particularly pronounced age-dependent patterns. Our results are consistent with this evidence and further reinforce the idea that relationships between the body, brain, and cognition in later life arise from interconnected, system-wide processes rather than isolated physiological functions (Picca et al., 2025).

From a clinical perspective, these results identify several modifiable risk factors, such as adiposity, sarcopenia, osteoporosis, impaired glucose metabolism, hypertension, and reduced pulmonary function, that may be targeted to preserve cognitive health. Conversely, protective factors, including lean muscle mass, bone integrity, lung capacity, and circulating IGF-1 levels, suggest biological pathways through which lifestyle interventions and preventive medicine could enhance brain health and cognitive resilience. High predictive power of body composition and BMD, rivalling that of conventional neuroimaging, reinforces the importance of integrating systemic physical health markers into cognitive risk assessment.

In summary, cognitive variations in later life reflect the intertwined influences of system-wide physiology and brain health. Recognising this dual contribution opens new avenues for prevention and treatment, emphasising that maintaining body health is inseparable from protecting brain health. Future research should build on these findings to develop integrated, clinically applicable models that capture the complexity of cognitive aging and translate them into effective strategies for preserving cognition across populations.

## LIMITATIONS

Our study is not without limitations. First, the cross-sectional design does not allow us to infer the temporal dynamics of cognitive and physical aging, which limits our ability to understand how these continuous processes unfold over time, particularly in older adults. Longitudinal research is therefore needed to capture developmental trajectories and to clarify potential mechanisms and directional relationships linking aging, cognition, and body and brain health.

Second, the predictive associations reported in this study, as well as the shared variance between body- and brain-derived predictors, should not be interpreted as evidence of mechanistic or causal pathways linking body physiology, brain characteristics, and cognition, because machine learning models do not establish the directionality of relationships. In this context, predictive performance indicates that body and brain measures contain information relevant to cognitive functioning but does not imply that these measures directly cause or determine cognitive outcomes. Because aging influences multiple physiological systems simultaneously, the observed overlap between body physiology, neuroimaging measures, and cognition may partly reflect shared age-related or broader health-related processes rather than direct biological mediation. Shared unmeasured factors, including genetic predisposition, early-life exposures, and socioeconomic influences, may also contribute to the observed associations. Future research using longitudinal designs and causal inference approaches, such as longitudinal mediation analyses or controlled animal studies, will be important for determining whether the relationships identified here reflect causal pathways or shared etiological influences.

Importantly, the findings may have limited generalizability to other populations due to cultural and societal characteristics specific to the UK Biobank sample (Fry et al., 2017; Keyes & Westreich, 2019; van Alten et al., 2024). For example, UK Biobank participants are generally healthier, more educated, and socioeconomically advantaged than the broader population, which may influence both physiological and cognitive measures and inflate observed associations (Bradley & Nichols, 2022; Fry et al., 2017; Lyall et al., 2022; Schoeler et al., 2023; van Alten et al., 2024). In addition, we did not stratify participants based on cognitive status, as cognition in older age spans a broad spectrum, with gradual transitions from normal functioning to severe impairment, and biomarkers of neurodegenerative pathology, such as amyloid or *tau*, were available only for a limited subset of participants. We also did not exclude individuals with neurological, psychiatric, or major systemic disorders, as these conditions become increasingly common with age, and excluding them would further increase healthy participant bias, reduce sample representativeness, and limit generalizability to the broader aging population. Consequently, such conditions may contribute additional shared variance across body- and brain-derived measures, and some observed age-related associations may partly reflect unmeasured pathological processes in addition to normative aging. Here, our aim was to characterize patterns of association in a large population-based cohort, and we encourage future studies to investigate brain–body–cognition relationships in more diverse samples with broader representation of sociodemographic backgrounds, educations, ethnicities, neurodegenerative biomarkers, and clinical conditions, although doing so may reduce the availability of certain features and subgroup sample sizes. Formally investigating the influence of these variables represents an important direction for future research. For example, commonality analysis could be used to quantify the extent to which relationships between cognition and these factors (e.g., cognition–socioeconomic status or cognition–education) overlap with brain and body markers, analogous to our analysis of cognition–age covariance.

Third, our study lacks cognitively demanding task-based functional MRI (tfMRI) measures, which may account for a substantial portion of condition-specific variance in particular cognitive domains (Tetereva et al., 2022). In the UK Biobank, tfMRI data have a smaller sample size and fewer features and come from the Hariri hammer task (Hariri et al., 2000). This task probes emotional reactivity, particularly amygdala responses, and is less cognitively demanding than other tfMRI paradigms, such as the *n*-back working memory task and the Face Name Associative Memory Exam (Kirchner, 1958; Tetereva et al., 2022, 2025; Tetereva & Pat, 2023).

Finally, cognitive tests within the UK Biobank battery differ from commonly used instruments such as the NIH Toolbox for Assessment of Neurological and Behavioral Function (Gershon et al., 2013) or the Wechsler Adult Intelligence Scale (Hartman, 2009), which are employed in other population-level studies (Casey et al., 2018; Elam & Van Essen, 2022; Poulton et al., 2015). This discrepancy makes it more difficult to generalize cognitive performance across cohorts.

## CONCLUSIONS

Taken together, our findings show that neuroimaging-based predictors of cognition substantially overlapped with the variance shared between system-wide physiology markers and cognition, accounting for over 85% of this shared variance. White matter integrity, body composition, and musculoskeletal health emerged as the strongest predictors. Having information about both the body and the brain allowed us to explain nearly all age-related variation in cognition. This convergence demonstrates that cognitive aging reflects the interplay between systemic physiology and brain structure and function, underscoring that cognition is embedded within whole-body physiology and is best understood as a multisystem process spanning body and brain.

## MATERIALS AND METHODS

We analysed multimodal data comprising 12 cognitive performance scores, 19 body phenotypes, and 81 brain phenotypes collected during the first imaging visit of the UK Biobank study (application 70132). As the aim of the study was to characterize population-level variability in physiological and cognitive functioning, participants were not excluded on the basis of neurological, psychiatric, or systemic medical conditions. The original sample used to derive the *g*-factor included 31,897 participants aged 46–83 years (mean = 64.55, SD = 7.66; 51.32% female; 91.3% British, 2.49% Irish, 3.3% other white backgrounds, <1% other ethnicities). The final sample in the commonality analysis included 25,346 participants aged 47–82 years (mean = 64.08, SD = 7.52; 53.18% female; 91.25% British, 2.5% Irish, 3.35% other white backgrounds, <1% other ethnicities).

### Data

#### General cognition factor

We quantified cognitive function as a latent general cognition factor (*g*-factor) derived from twelve cognitive performance scores (see Supplementary Table S1 for a list of cognitive tests and scores included in the study) using exploratory structural equation modelling within confirmatory factor analysis (ESEM-within-CFA, https://mateuspsi.github.io/esemComp/articles/esem-within-cfa.html). To reduce skewness, we applied a *log*-transformation to the mean time required to correctly identify matches in the Reaction Time test, to the time needed to complete numeric and alphabetic trails in the Trail Making test, and a *log(x+1)* transformation to the number of incorrect matches in the six-pair version of the Pairs Matching task (Fawns-Ritchie & Deary, 2020; Lyall et al., 2016). For the Symbol Digit Substitution test, the accuracy rate was computed as the ratio of correctly matched symbol–digit pairs to the total number attempted, to account for spurious correct responses arising from rapid guessing. This approach mitigates the inflation of scores that may occur when participants respond quickly without accuracy, since occasional correct matches can occur by chance. Overall, cognitive performance measures encompassed five cognitive domains: executive functions, fluid intelligence, reaction time and processing speed, memory, and crystallised cognitive ability (Fig. 1) (Fawns-Ritchie & Deary, 2020).

We derived the *g*-factor using a hierarchical model structure based on a set of domain-specific factors. To evaluate model fit, we used the Comparative Fit Index, Tucker–Lewis Index, Root Mean Square Error of Approximation, Bayesian Information Criterion, and Standardized Root Mean Square Residuals (Supplementary Tables S3 and S4) (Buianova et al., 2026; Ritchie et al., 2016; Tucker-Drob et al., 2014).

#### Body phenotypes

As measures of physical health, we used 19 body phenotypes covering body composition, cardiovascular, pulmonary, renal, hepatic, immune, metabolic, and musculoskeletal systems (Supplementary Table S2). Body composition was assessed using magnetic resonance imaging (MRI), bioelectrical impedance analysis, and dual-energy X-ray absorptiometry (DXA). Bone size, mineral content, and density were measured by DXA. Cardiovascular health was characterized through carotid ultrasound, arterial stiffness indices, pulse wave analysis, heart MRI for left ventricular size and function, and 12-lead resting electrocardiogram (ECG). Renal and hepatic structure was examined using MRI of the kidney, liver, and abdominal organs. Additional assessments included blood-based markers of renal and hepatic functions, immune and metabolic health, and hearing tests. Variables with more than 60% missing data, including rheumatoid factor, blood oestradiol, microalbumin in urine, cardiorespiratory fitness, ECG during exercise, kidney distance, total adipose tissue volume, and total lean tissue volume, were excluded from the analyses.

#### Neuroimaging

We derived brain markers of cognition from 42 diffusion-weighted MRI (dwMRI), 18 resting-state functional MRI (rsMRI), and 21 structural MRI (sMRI) neuroimaging phenotypes (Supplementary Table S4). dwMRI assesses white matter microstructure, including fibre orientation and density, by measuring the directionality of water diffusion within myelinated axons (Basser et al., 1994). rsMRI captures spontaneous blood-oxygenation-level–dependent (BOLD) signal fluctuations, enabling the estimation of functional connectivity both between cortical and subcortical regions and across large-scale networks (M. H. Lee et al., 2013). sMRI leverages T1- and T2-weighted MRI sequences to quantify brain morphology like cortical thickness, grey and white matter volumes, and surface area (Symms et al., 2004; Wattjes, 2011).

##### Diffusion-weighted MRI (dwMRI)

The dwMRI brain phenotypes included diffusion tensor imaging and neurite orientation dispersion and density imaging metrics, such as fractional anisotropy (FA), diffusion tensor mode, mean diffusivity, and the three eigenvalues of the diffusion tensor, as well as intracellular volume fraction, isotropic or free water volume fraction, and the orientation dispersion index (Supplementary Table S4). These metrics were extracted for 48 white matter tracts using tract-based spatial statistics and for 27 white matter tracts using probabilistic tractography. Additionally, we used structural connectome data (Mansour et al., 2023), represented as matrices containing quantitative metrics, including streamline count, fibre bundle capacity (from spherical-deconvolution informed filtering of tractograms), mean streamline length, and mean FA, for each node pair, derived from six different combinations of cortical and subcortical atlases (Mansour et al., 2023): the Desikan–Killiany (aparc) cortical atlas (Desikan et al., 2006) + Melbourne Subcortical Atlas scale I (MSA-I) (Tian et al., 2020), the Destrieux (aparc.a2009s) cortical atlas (Destrieux et al., 2010) + MSA-I (Tian et al., 2020), the Glasser cortical atlas (Glasser et al., 2016) + MSA-I (Tian et al., 2020), the Glasser cortical atlas (Glasser et al., 2016) + MSA-IV (Tian et al., 2020), the Schaefer atlas for 200 cortical regions (7 networks) (Schaefer et al., 2018; Yeo et al., 2011) + MSA-I (Tian et al., 2020), and the Schaefer atlas for 500 cortical regions (7 networks) (Schaefer et al., 2018; Yeo et al., 2011) + MSA-IV (Tian et al., 2020).

##### Resting-state functional MRI (rsMRI)

The rsMRI brain phenotypes included full and partial correlation matrices for 21 and 55 components derived from independent component analysis (ICA), as well as functional connectomes for the same six cortical–subcortical atlas combinations used for the dwMRI structural connectomes. For ICA-based networks, we additionally used network amplitudes. To construct functional connectomes for parcellated data, we combined BOLD time series from cortical and subcortical parcellations and computed both full and partial correlation matrices using the *ConnectivityMeasure* function in *Nilearn* (Buianova et al., 2026).

##### Structural MRI (sMRI)

The sMRI brain phenotypes from T1- and T2-weighted MRI included regional and subcortical grey matter volumes and mean intensity; cortical and white matter surface area, thickness, and volume; grey–white matter contrast intensity; pial surface area; and total volumes of white matter hyperintensities from FSL FAST and FSL FIRST, FreeSurfer’s automated subcortical volumetric segmentation (ASEG), *ex-vivo* Brodmann Area Maps, Destrieux (a2009s) parcellation, Desikan–Killiany–Tourville parcellation, Desikan–Killiany parcellation, and subcortical volumetric subsegmentation (Buianova et al., 2026).

### Data Analysis

#### Machine learning

We applied machine learning to predict the *g*-factor separately from each body and brain phenotype and then combined these predictions into multimodal biomarkers using a stacking approach. For each first-level model, *g*-factor scores were matched to body and neuroimaging features using participant identifiers, and only individuals with complete data for all variables were retained. Prior to stacking, predicted cognitive scores generated from each body and/or neuroimaging phenotype were combined and used as input features for the second-level models. Because different first-level models were based on partially overlapping participant subsets, some missing values were present in the combined feature sets and were retained during stacking. Predicted scores from the stacked models were subsequently used as explanatory variables in the commonality analyses. For these analyses, predicted and observed *g*-factor values were matched using participant identifiers, and only participants with complete data across the explanatory variables included in a given model were retained, with only out-of-sample predictions from the test sets used to ensure independence in the commonality analyses.

Performance of machine learning models was estimated using nested cross-validation with five outer folds and ten inner folds, with hyperparameters tuned exclusively within the inner loops to avoid optimization bias. In each outer fold, 20% of the data served as the held-out test set, while the remaining 80% formed the training set. This training set was further partitioned into ten inner folds for hyperparameter optimisation through cross-validation. Models were trained on nine folds and validated on the remaining one, iterating across all ten combinations. Using five outer folds and ten inner folds offered an optimal balance between computational efficiency, robust hyperparameter tuning, sufficient sample size for model training, and adequate test set sizes for model evaluation.

We selected the hyperparameter configuration that achieved the lowest mean squared error (*MSE*) across the inner-fold validation sets. Using this configuration, we retrained the model on the full outer-fold training set and evaluated on the corresponding outer-fold test set, from which predicted values were obtained. Performance was quantified using Pearson r, the coefficient of determination (*R*^2^), the mean absolute error (*MAE*), and the *MSE*. This procedure was repeated across all five outer folds, ensuring that every data point contributed to both training and testing, but never simultaneously.

Before building machine learning models, all variables were standardized using the mean and standard deviation from the training set, with these parameters applied to the corresponding test set within each outer fold, to avoid data leakage. Standardisation was carried out at four stages: cognitive performance scores were standardized before ESEM; neuroimaging data were standardized before regressing out modality-specific confounds from brain features; body phenotypes were standardized before first-level model training; and first-level predictions together with observed targets were standardized before building stacked models.

Before deriving neural markers of cognition, we corrected each set of brain phenotypes for common and modality-specific confounds, which account for scanner position, acquisition and reconstruction parameters, motion, image quality metrics, and modality-specific artefacts, following the validated UK Biobank imaging confound pipeline developed by Alfaro-Almagro and colleagues (2021) (see Supplementary Table S4 for the complete list of neuroimaging confounds). We did not model the assessment site separately, as imaging data collection was harmonized across acquisition sites, and the confound set already accounts for variance components that may overlap with site-related effects. Confounds with pairwise correlations below *r* = 0.7 were retained to mitigate multicollinearity. After standardising neuroimaging features and confounds, we applied linear regression to obtain residuals for each brain phenotype, which were then used as input features for the machine learning models (Buianova et al., 2026). Throughout the analysis, both base and stacked models were trained solely on participants from the outer-fold training set and evaluated on the corresponding outer-fold test set, ensuring a strict separation between training and test data.

To derive markers of cognition, we used a two-step machine learning approach. First, we trained individual *XGBoost* models for each brain or body phenotype to obtain phenotype-specific predictions of the *g*-factor. *XGBoost* is a gradient boosting method that sequentially builds decision trees, where each tree corrects the residual errors of the previous ensemble (Chen & Guestrin, 2016). It incorporates regularisation of model complexity and feature weights, improving generalizability and reducing overfitting. *XGBoost* is well suited to high-dimensional data and nonlinear relationships, and, for tabular data, it often outperforms many deep learning algorithms (Shwartz-Ziv & Armon, 2022).

In the second step, we applied multimodal stacking to integrate these predictions across phenotypes. Here, first-level predictions (i.e., phenotype-specific *g*-factor estimates) were used as input features for a second-level model, which combined information across modalities to generate a final prediction (Buianova et al., 2026; Tetereva et al., 2025; Wolpert, 1992). We constructed six stacked models: one for all body phenotypes, one combining all brain phenotypes, three modality-specific brain models (dwMRI, rsMRI, sMRI) combining neuroimaging phenotypes within each modality, and one combined brain–body model integrating both brain and body phenotypes. For stacking, we used *Random Forest*, which aggregates predictions from multiple decision trees trained on bootstrap samples (Breiman, 2001; Schonlau & Zou, 2020). At each split, a random subset of predictors is considered, which decorrelates trees and improves stability. The model captures nonlinear effects and interactions, is robust to multicollinearity, and accommodates missing values that arise during integration of phenotype-specific first-level predictions via surrogate splits. Hyperparameter tuning involved up to twelve parameters for *XGBoost* and two for *Random Forest* (see Supplementary Table S20). For feature sets exceeding 30,000 variables, optimization used *RandomizedSearchCV*, while smaller sets were tuned using *GridSearchCV*.

To assess the statistical significance of the models, we applied bootstrapping to the predicted and observed *g*-factor values aggregated across all five outer-fold test sets. We generated an empirical distribution of Pearson correlation coefficients by resampling the data with replacement for 5,000 iterations and derived 95% confidence intervals (CIs) from this distribution. Model performance was considered statistically significant when the 95% CI excluded zero, suggesting that the observed relationships were unlikely to occur by chance.

Additionally, we benchmarked the predictive performance of the top-performing body phenotype against the other body phenotypes as well as the composite body marker against unimodal and composite brain markers. Comparisons included (a) the top-performing body phenotype versus the remaining individual body phenotypes, (b) composite body marker versus individual body and brain phenotypes, unimodal and composite brain markers, and a body+brain marker, and (c) the top-performing brain phenotype versus individual body and brain phenotypes, unimodal and composite brain markers, composite body marker, and a body+brain marker. To assess the difference in predictive performance, we compared four performance metrics: Pearson *r*, *R*^2^, *MSE*, and *MAE*.

#### Feature importance

To facilitate interpretation of feature importance in the machine learning models, we applied the Haufe transformation (Chen et al., 2023; Chopra et al., 2024; Haufe et al., 2014; Tian & Zalesky, 2021) to model predictions pooled across the five outer-fold test sets. The Haufe transformation converts predictive model weights into feature-wise associations with the predicted outcome while accounting for the covariance structure of the data (Haufe et al., 2014), thereby enabling identification of features most relevant to model prediction. Within this framework, to explain how each of the 19 body phenotypes was related to predictions generated by the stacked model combining all body phenotypes, we computed correlations between the composite body marker and the prediction derived from each body phenotype. Bonferroni correction for multiple comparisons was applied across these 19 phenotype-level tests. Similarly, to explain how each of the 317 body physiology measures across different body phenotypes was related to the predictions of the stacked model, we computed correlations between the composite body marker and each body physiology measure. Here, the Bonferroni correction was applied across these 317 feature-level tests. Lastly, to characterize which neuroimaging features contributed most strongly to the predictions made by the stacked model combining all brain phenotypes, we computed correlations between the composite brain marker and neuroimaging features from the top-performing brain phenotype within each neuroimaging modality. These analyses were intended for descriptive interpretation of model feature importance rather than formal mass-univariate statistical inference.

### Commonality analysis

We employed commonality analysis (Nimon et al., 2008; Viswesvaran, 1998) to quantify (a) the extent to which the cognition–body relationship overlapped with unimodal and composite brain markers, and (b) the extent to which the cognition–age relationship overlapped with composite body and/or brain markers. This involved fitting a series of linear regression models in which the observed *g*-factor, pooled across five outer-fold test sets, served as the response variable, and predicted *g*-factors based on body or brain, or age (also pooled across folds) were included as explanatory variables. To ensure independence, only outer-fold test sets were used, such that the predicted *g*-factors (predictors) were independent of the data used to train the models that generated them (outer-fold training sets). The regression models included:

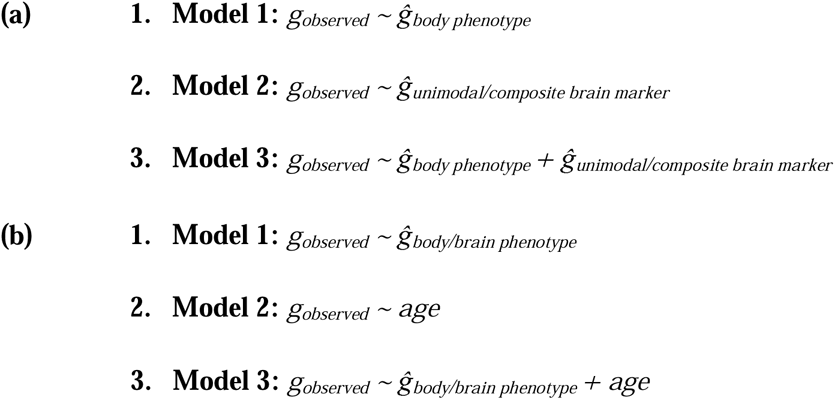

This approach enabled us to estimate the variance in the observed *g*-factor accounted for by the predicted *g*-factor based on body or brain, or age. To quantify overlap in variance between (a) brain markers and the cognition–body relationship and (b) body and brain markers and the cognition–age relationship, we decomposed the variance of the observed *g*-factor into components explained uniquely or commonly by each pair of explanatory variables. We then expressed these overlaps in variance for each explanatory variable as a percentage ratio, where the numerator corresponded to the variance commonly explained by the two explanatory variables and the denominator corresponded to the total variance in observed *g*-factor explained by the explanatory variable of interest, computed as:

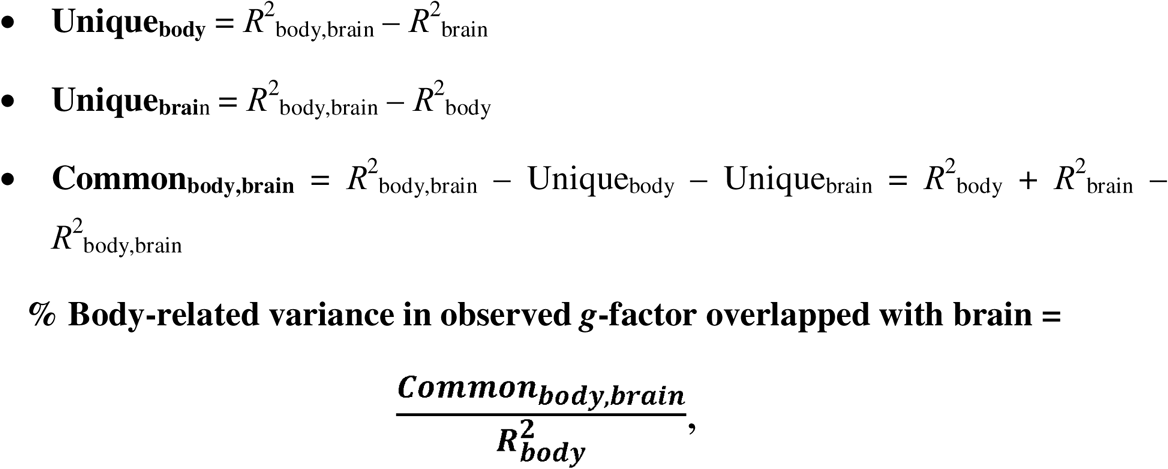

where *R*^2^_body_ = Common_body,brain_ + Unique_body_.

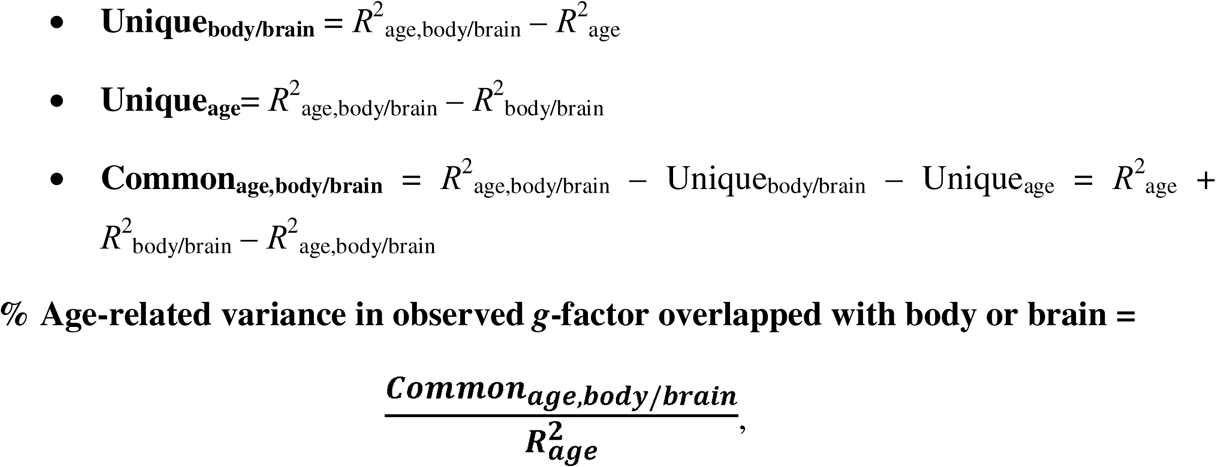

where *R*^2^_age_ = Common_age,body/brain_ + Unique_age_.

Here, *Unique* denotes the variance in cognition uniquely overlapped with one of the explanatory variables; *Common* denotes the variance commonly overlapped with two explanatory variables; and *R*^2^ represents the total variance in cognition attributed to each explanatory variable – or to pairs of variables – in independent linear regression models (Nimon et al., 2008; Viswesvaran, 1998).

To quantify how composite body and brain markers commonly overlapped with age-related variation in cognition, we extended the commonality analysis to a three-explanatory-variable model:

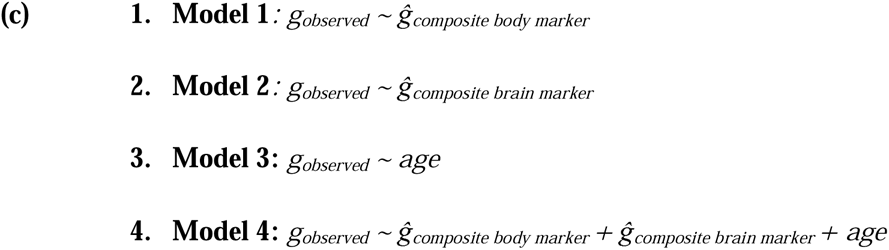

We decomposed the variance of the observed *g*-factor into components unique to age, body, and brain, as well as components common to each pair of explanatory variables and to all three explanatory variables together. This decomposition enabled us to estimate the proportion of age-related cognitive variance that can be uniquely or commonly overlapped with body and brain markers.

We then expressed the overlaps as a percentage ratio, where the numerator corresponded to the variance in the observed *g*-factor commonly explained by the explanatory variable of interest and age (with or without the third explanatory variable), and the denominator corresponded to the total variance in observed *g*-factor explained by age:

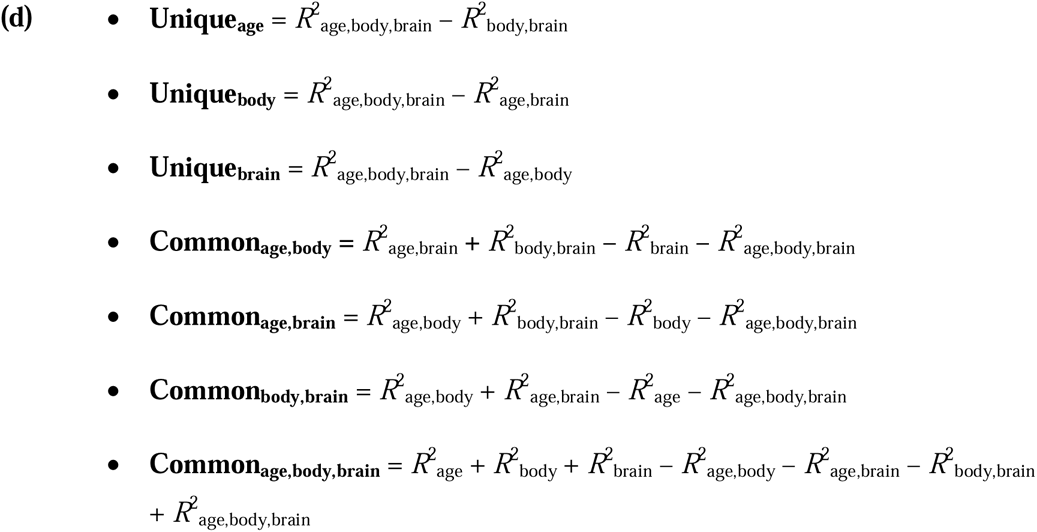

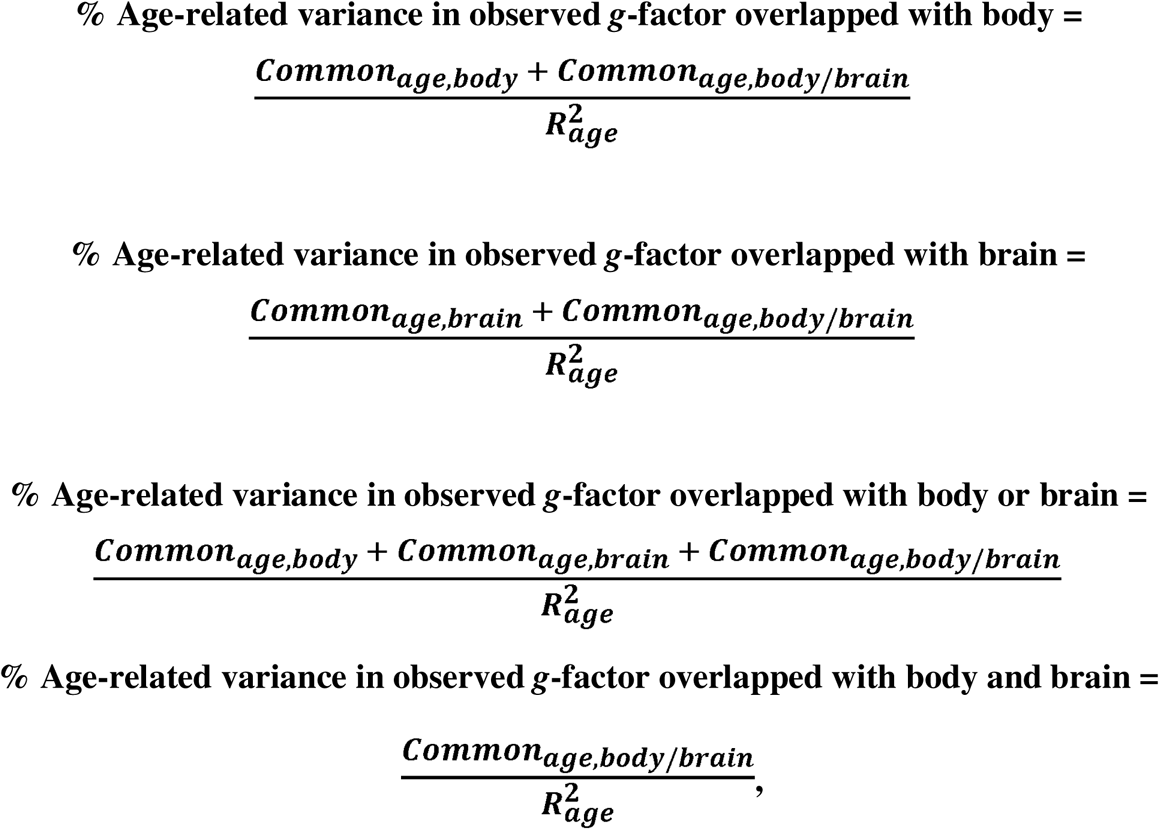

where *R*^2^_age_ = Unique_age_ + Common_age,body_+ Common_age,brain_+ Common_age,body,brain_.

### Mediation Analysis

In addition to the commonality analyses, we conducted formal mediation analyses for completeness (Rijnhart et al., 2021). These analyses examined the extent to which brain markers mediated the relationship between cognition and body measures, as well as the extent to which body physiology and brain markers jointly mediated the relationship between cognition and age. Notably, mediation analyses rely on stronger assumptions about directional causality. Moreover, mediation analysis addresses questions about potential causal pathways (“how much of an effect is transmitted through a mediator”), whereas commonality analysis addresses questions about shared variance (“how much variance is explained jointly”). Results of the mediation analyses are presented in Supplementary Table S21 and Supplementary Figures S1 and S2.

## Data Availability

The data used in this study are subject to restrictions. Access requests should be directed to the UK Biobank (https://www.ukbiobank.ac.uk/).

## Code Availability

All analysis code is available at https://github.com/HAM-lab-Otago-University/UKBiobank-Brain-Body.

## Acknowledgements

This research has been conducted using the UK Biobank Resource under Application Number 70132. N.P. was supported by Health Research Council of New Zealand (grant numbers 21/618 and 24/838), by Neurological Foundation of New Zealand (grant number 2350 PRG), and by the Ministry of Business, Innovation and Employment (grant numbers UOA2421 and RTVU2403). I.B. was supported by the University of Otago.

