## Supplemental Figures for "Exploring the Link Between Body Physiology and Cognition: The Role of the Brain and Aging"

### Supplementary Information

Irina Buianova, MSc, Narun Pat, PhD

Department of Psychology, University of Otago, Dunedin, New Zealand

**Figure S1**. Mediation analysis examining the indirect association between body physiology (composite body marker) and cognitive functioning (*g*-factor) via brain neuroimaging phenotypes stacked across three (A), within dwMRI (B), rsMRI (C), and sMRI (D) modalities

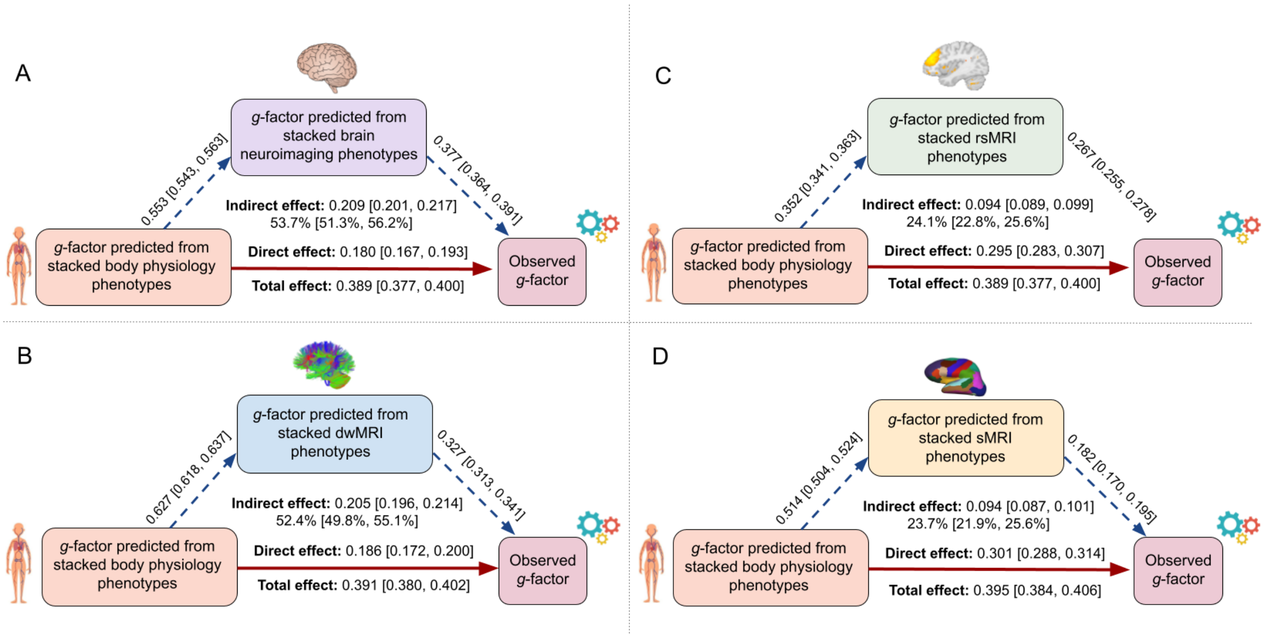


The model treated the composite body marker (out-of-sample *g*-factor predictions derived from stacked body physiology phenotypes) as the independent variable, the composite brain marker (out-of-sample predictions derived from neuroimaging phenotypes stacked within and across MRI modalities) as the mediator, and the observed *g*-factor as the dependent variable. All variables were standardized (*z*-scored); therefore, all reported coefficients are standardized. Parentheses indicate 95% confidence intervals based on 5,000 bootstrap resamples. Mediation analysis was conducted using *lavaan* (version 0.6-21). All effects were statistically significant at $p < 0.001$ (Supplementary Table S21).

**Figure S2.** Results of the mediation analysis examining indirect associations between age and cognitive functioning (*g*-factor) via body physiology (composite body marker) and brain neuroimaging phenotypes stacked across three MRI modalities (composite brain marker)

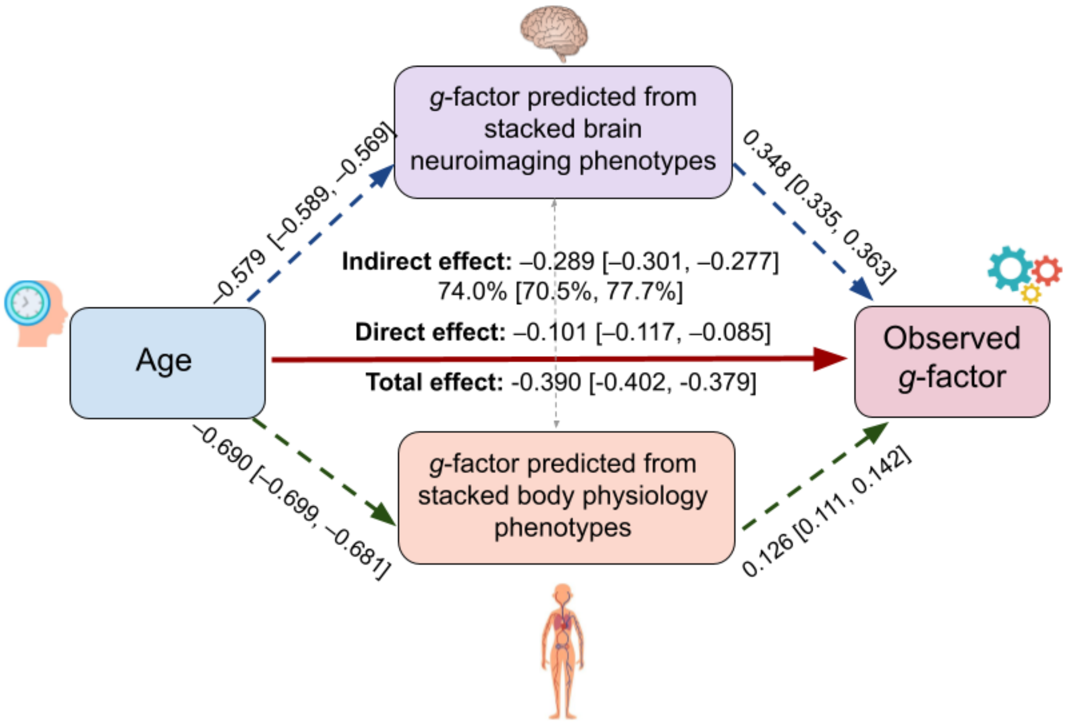


The model treated age as the independent variable, the composite body marker (out-of-sample *g*-factor predictions derived from stacked body physiology phenotypes) and the composite brain marker (out-of-sample predictions derived from neuroimaging phenotypes stacked across all MRI modalities) as parallel mediators, and the observed *g*-factor as the dependent variable. The indirect effect reported in the figure represents the combined indirect effect of both mediators.

The indirect effect through the composite brain marker was –0.202 (95% CI [–0.210, –0.193]), corresponding to 51.7% (95% CI [49.1%, 54.4%]) of the total effect. The indirect effect through the composite body physiology marker was –0.087 (95% CI [–0.098, –0.077]), corresponding to 22.3% (95% CI [19.6%, 25.2%]) of the total effect. All variables were standardized (*z*-scored); therefore, all reported coefficients are standardized. Parentheses indicate 95% confidence intervals based on 5,000 bootstrap resamples. Mediation analysis was conducted using *lavaan* (version 0.6-21). All effects were statistically significant at $p < 0.001$. (Supplementary Table S21).
